# Adverse Psychosocial Trajectory in Bipolar Disorder: Novel Genetic Links to ADHD and Anxiety

**DOI:** 10.1101/2025.05.17.25327825

**Authors:** Tracey van der Veen, Nick Bass, Andrew McQuillin

**Author notes:** Correspondence to: Tracey van der Veen, Molecular Psychiatry Laboratory, Division of Psychiatry, University College London, London, WC1E 6BT, UK.

## Abstract

**Background:** Bipolar disorder (BD) factor models offer limited dimensional understanding due to incomplete integration of psychosocial deficits, long-term outcomes, and transdiagnostic genetics, thus restricting personalised interventions. This study aimed to provide a holistic understanding of BD psychopathology, overcoming this limitation.

**Methods:** Exploratory Factor Analysis of 77 OPCRIT items revealed four psychopathological dimensions, and Confirmatory Factor Analysis validated a 20-item, four-factor BD model. Polygenic Risk Scores for five relevant disorders were calculated, and Structural Equation Modelling analysed the genetic contributions to this dimensional model. The study applied Inverse Probability Weighting to address biases in a sample of 4992 participants.

**Results:** Confirmatory Factor Analysis revealed a novel Adverse Psychosocial Trajectory (APT) dimension, characterised by the co-occurrence of premorbid deficits and poorer long-term outcomes in individuals with BD. Structural Equation Modelling further showed distinct patterns of genetic liability: BD PRS for mania, Schizophrenia (SCZ) PRS for psychosis, and Major Depressive Disorder (MDD) PRS for depression. Notably, the APT dimension exhibited a positive association with Attention-Deficit/Hyperactivity Disorder (ADHD) and anxiety PRSs, and an inverse relationship with BD PRS.

**Conclusions:** This study offers a novel and clinically relevant dimensional model of BD by identifying the APT dimension, which uniquely integrates crucial premorbid psychosocial factors and outcomes. The identified direct genetic link between ADHD and anxiety with APT (a trajectory associated with poorer BD outcomes) provides important new insight into a challenging illness course. This potentially enables earlier identification and facilitates targeted interventions to improve long-term psychosocial outcomes and overall quality of life in BD.

## Introduction

### Limitations of Categorical Diagnosis

Bipolar disorder (BD) shows diverse outcomes influenced by genetics beyond current subtyping (BD1, BD2). Traditional classifications often overlook the critical impact of premorbid psychosocial factors on long-term outcomes. While course specifiers aim to improve treatment alignment (Regier, Kuhl, & Kupfer, 2013; OConnell & Coombes, 2021), this study proposes a novel dimensional approach for a more nuanced understanding of BDs inherent heterogeneity beyond categorical diagnoses.

### Bipolar Disorder: A Symptom Continuum

Many BD patients experience continuous symptoms beyond discrete episodes: cognitive deficits in remission are reported, with prevalence as high as 70% (Pavlova et al., 2017; Sanches et al., 2015; Mignogna & Goes, 2024); 20-50% experience inter-episodic symptoms (Grunze & Born, 2020), highlighting limitations of episodic models. Even during euthymia, executive dysfunction and anxiety persist, indicating vulnerability (Altshuler et al., 2004; Robinson et al., 2006). Personality traits also influence BDs onset, progression, and course (Sparding, Pålsson, Joas, Hansen, & Landén, 2017).

### Dimensional Frameworks in Bipolar Disorder

Dimensional approaches dissect BDs heterogeneity, allowing researchers to identify potentially more genetically similar subgroups based on specific symptom profiles. Acknowledging this heterogeneity, research increasingly focuses on genetic differences within more homogeneous subgroups (Schulze, 2010) to understand genetic contributions to diverse presentations. While specific BD course specifiers show familiality (Saunders, Scott, McInnis, & Burmeister, 2008), and genetic liabilities for subphenotypes are being identified, single regression models can complicate interpretation (Allardyce, McCreadie, Morrison, & van Os, 2007; Ruderfer et al., 2014; Reininghaus et al. (2016); Qiu, Akiskal, Kelsoe, & Greenwood, 2017; Allardyce et al., 2018; Bipolar Disorder and Schizophrenia Working Group of the Psychiatric Genomics Consortium et al., 2018; Grigoroiu-Serbanescu et al., 2020; Coombes et al., 2020; Richards et al., 2022; Allardyce et al., 2023; Grigoroiu-Serbanescu et al., 2024; Song et al., 2024; Lawrence, Breunig, Foote, Tallis, & Grotzinger, 2024). A dimensional framework offers a powerful alternative by examining psychopathology along continuous axes, enabling nuanced analysis of specifier interrelations and combined genetic liabilities for a holistic understanding of BD heterogeneity.

### Impact of Psychosocial Deficits on Bipolar Disorder

Cognitive and psychosocial deficits, not fully recognised specifiers (DSM-5; American Psychiatric Association, 2013; Regier et al., 2013), contribute to BD variability and impair quality of life, even during mood stability (Sanches, Bauer, Galvez, Zunta-Soares, & Soares, 2015; Miskowiak, Hansen, Mariegaard, & Kessing, 2023). These deficits exist on a spectrum, negatively impacting relationships and productivity, often leading to social withdrawal (Judd et al., 2005; Bennett et al., 2019) and affecting 30-60% of adults with BD (Bennett et al., 2019). Early onset of these deficits links to worse outcomes like anxiety, substance use, and suicidality, with increased childhood risk (Carballo et al., 2020; Rohde, Lewinsohn, Seeley, Klein, Andrews, & Small, 2007). Recognising genetic predisposition could potentially reduce diagnostic delays (Scott et al., 2022) and suicide rates in BD (Dong et al., 2019). Examining these deficits within a broader psychopathological spectrum may also clarify connections to other disorders. While research on psychosis (Allardyce, McCreadie, Morrison, & van Os, 2007) explored premorbid risk factors, their specific impact on long-term BD outcomes remains less understood.

### Genetic Contributions and Polygenic Risk Scores

Genetic factors substantially contribute to BD comorbidity (Léda-Rêgo et al., 2024), with approximately 35-65% of individuals with BD meeting criteria for another psychiatric condition (Salvi, Ribuoli, Servasi, Orsolini, & Volpe, 2021), indicating complex psychopathology interplay. This high comorbidity suggests single-disorder analyses might miss critical genetic factors contributing to this broader spectrum of co-occurring conditions. Symptoms often begin early and persist, worsened by environmental factors (Aas et al., 2020; Park, Shekhtman, & Kelsoe, 2020; Schoeler et al., 2019; Baldwin et al., 2021; Yao, van der Veen, Thygesen, Bass, & McQuillin, 2023).

Polygenic risk scores (PRSs) are valuable tools for investigating the genetic basis of BD heterogeneity and comorbidities (Wang, Tsuo, Kanai, Neale, & Martin, 2022; Mitchell et al., 2023). Analysing symptom clusters may reveal stronger genetic associations than isolated disorder analyses. For example, higher SCZ PRS is linked to mood-incongruent psychotic symptoms and earlier BD onset (Allardyce et al., 2018; Grigoroiu-Serbanescu et al., 2024). Higher ADHD and anxiety risk correlates with rapid cycling (Coryell et al., 2003; Kupka et al., 2005; Coombes et al., 2020).

ADHD increases multimorbidity risk, worsening symptom severity and functional impairment (Du Rietz et al., 2018; Vannucchi et al., 2019; Mignogna & Goes, 2024). Polygenic ADHD burden has been linked to earlier BD onset and lithium resistance, while lithium response can be influenced by family history and absence of anxiety or rapid cycling (Dunner & Fieve, 1974; van Hulzen et al., 2017; Grigoroiu-Serbanescu et al., 2020; Coombes, Millischer, Batzler, Larrabee, Hou, Papiol, Heilbronner, Adli, Akiyama, Akula et al., 2021; Lin et al., 2021; Patel et al., 2024). Factor analysis can simplify complex relationships between symptoms and disorders, revealing underlying factors and their genetic contributions within a spectrum framework.

### Introducing Adverse Psychosocial Trajectory (APT)

Building on prior BD modelling using OPCRIT items (Allardyce et al., 2023), this study introduces a novel four-factor model. By combining OPCRIT items and PRS, we identified an Adverse Psychosocial Trajectory (APT) dimension, demonstrating correlations between premorbid psychosocial deficits and adverse BD outcomes, with shared genetic burdens for ADHD and anxiety prominently associated with APT, thus emphasising its role and genetic links for advancing BD understanding, classification, and intervention.

## Methods

### Study Design and Materials

This cross-sectional study examined relationships between clinical symptoms and genotypes linked to bipolar disorder, schizophrenia, depression, anxiety, and ADHD. All participants provided written informed consent. Research psychologists or psychiatrists conducted semi-structured interviews with bipolar disorder patients using the well-validated 90-item OPCRIT (Williams, Farmer, Ackenheil, Kaufmann, & McGuffin, 1996), which assesses premorbid functioning (personality, social and occupational adjustment) and longitudinal course (inter-episode patterns and chronicity), central aspects of our novel Adverse Psychosocial Trajectory (APT) dimension. This systematic approach enabled investigation of the interrelationship and genetic underpinnings of these crucial factors.

### Participants

Genotype data from 4992 individuals was analysed post-quality control. Clinical symptoms were assessed in 2590 individuals with DSM-IV bipolar disorder diagnoses (BD1, BD2, Bipolar Disorder Not Otherwise Specified [BD-NOS], or schizoaffective bipolar type (SZA)). Age of onset was determined by the first manic, mixed, or major depressive episode. Healthy controls were research volunteers screened for the absence of mental illness (and several unscreened random blood donors).

### Polygenic Risk Score Profiling

Genotyping was performed at the Broad Institute using Affymetrix Gene Chip 500k, Illumina PsychArray, and Illumina Global Screening Array (GSA). Post-QC variants and individuals were retained. The median number of post-QC variants was 3,164,648, used for polygenic risk score (PRS) analysis with imputed dosage files, genome-wide summary statistics for each psychiatric disorder, and a linkage disequilibrium (LD) reference panel. Continuous-shrinkage PRS (PRS-CS-auto) software (Ge, Chen, Ni, Feng, & Smoller, 2019), a Bayesian framework with default automatic shrinkage, calculated five trait PRSs per participant using summary statistics from the largest, most recent GWAS for BD (OConnell et al., 2025; *N*=840,309), depression (Howard et al., 2019; *N*=500,199), schizophrenia (Trubetskoy et al., 2022; *N*=130,644), anxiety disorder (Purves et al., 2020; *N*=114,091), and ADHD (Demontis et al., 2023; *N*=225,534). These GWASs were selected for their size and recency. The BD GWAS had the largest sample size. SCZ, MDD, ADHD, and anxiety frequently co-occur with BD and share genetic risk factors, making their PRS relevant for investigating BDs transdiagnostic genetic architecture. See Supplementary Sections S1-S15 for more detailed methods information.

### Controlling for Observational Study Biases

To mitigate confounders and ascertainment bias, Inverse Probability Weighting (IPW) using propensity scores (Robins, 1986; van der Wal & Geskus, 2011) was applied to cases and controls. This method adjusted for imbalances between groups and accounted for varying symptom severity in cases, ensuring more reliable genetic risk comparisons by reducing false associations from study inclusion. Stabilised weights were used. Additional covariates of age at interview, sex, and genotyping array were included with IPW propensity scores, with population stratification further accounted for by regressing out the first 10 ancestry-specific principal components (S14-15). Sensitivity analyses further confirmed result robustness (Mitchell et al., 2023; Wang et al., 2022).

### Statistical analyses

The primary aim of this study was to investigate the underlying structure of clinical symptoms in our bipolar disorder sample using factor analysis. Given the comprehensive nature of the OPCRIT instrument and our interest in identifying latent dimensions of psychopathology, we employed both Exploratory Factor Analysis (EFA) to uncover potential factors and Confirmatory Factor Analysis (CFA) to test the replicability and fit of a predefined factor structure. This approach allows for a data-driven exploration of symptom co-occurrence followed by a rigorous assessment of the identified factors.

Data for 77 clinical symptoms with adequate sample sizes were included for analysis (S7). Balanced clinical sample splits were used for exploratory and confirmatory phases. Items with zero-or near zero-variance (low frequency) were removed for latent factor modelling convergency. Low-frequency items were subsequently analysed separately via regression. Redundancy (multi-collinearity) was assessed using hetcor in the polycor (Olsson, 1979; Fox, 2022) R package. Highly correlated items (0.7+) were reviewed, retaining the least missing and most clinically relevant. Missingness was low (8%), with 92% data present in 77 items (var_miss in Naniar (Tierney, Cook, & Lumley, 2023)).

Missing at Random (MAR) was indicated by large *p*-values in chi-square tests (missing_compare in finalfit (finalfit team, 2023)). Imputation was avoided due to potential overfitting (Enders, 2022). The createDataPartition in Caret (Kuhn, 2008) created balanced 60/40 splits stratified by sex within subtypes.

Initial exploratory factor analysis (EFA) identified common factors from 77 OPCRIT items using efa in Lavaan (Rosseel, 2012). Higher factor loadings indicated stronger associations (S6-7). EFA included 1554 BD patients (60% calibration subsample) (S7). Four factors were retained based on parallel analysis (fa.parallel in psych (Revelle, 2023)), Scree Plot (S5), lower RMSEA, and clinical relevance. Ordinal categorical items were analysed with WLSMV estimator; *post hoc* Maximum Likelihood (ML) comparison showed no differences. Geomin rotation allowed factor correlation, while loadings remained uncorrelated (S6).

Confirmatory factor analysis, cfa in Lavaan, tested the reproducibility of factor loadings for 20 items within a four-factor framework (1036 validation sample). Twenty symptoms were selected based on a median 0.6+ factor loading and literature relevance. Model fit was assessed with EFA indices. No multicollinearity was suggested by Eigenvalue indices. Linearity assumption was checked via factor scores (lavPredict), and lavaanPlot was used for visualising the path diagrams (Lishinski, 2024) (Figure 1, S8).

**Figure 1.**
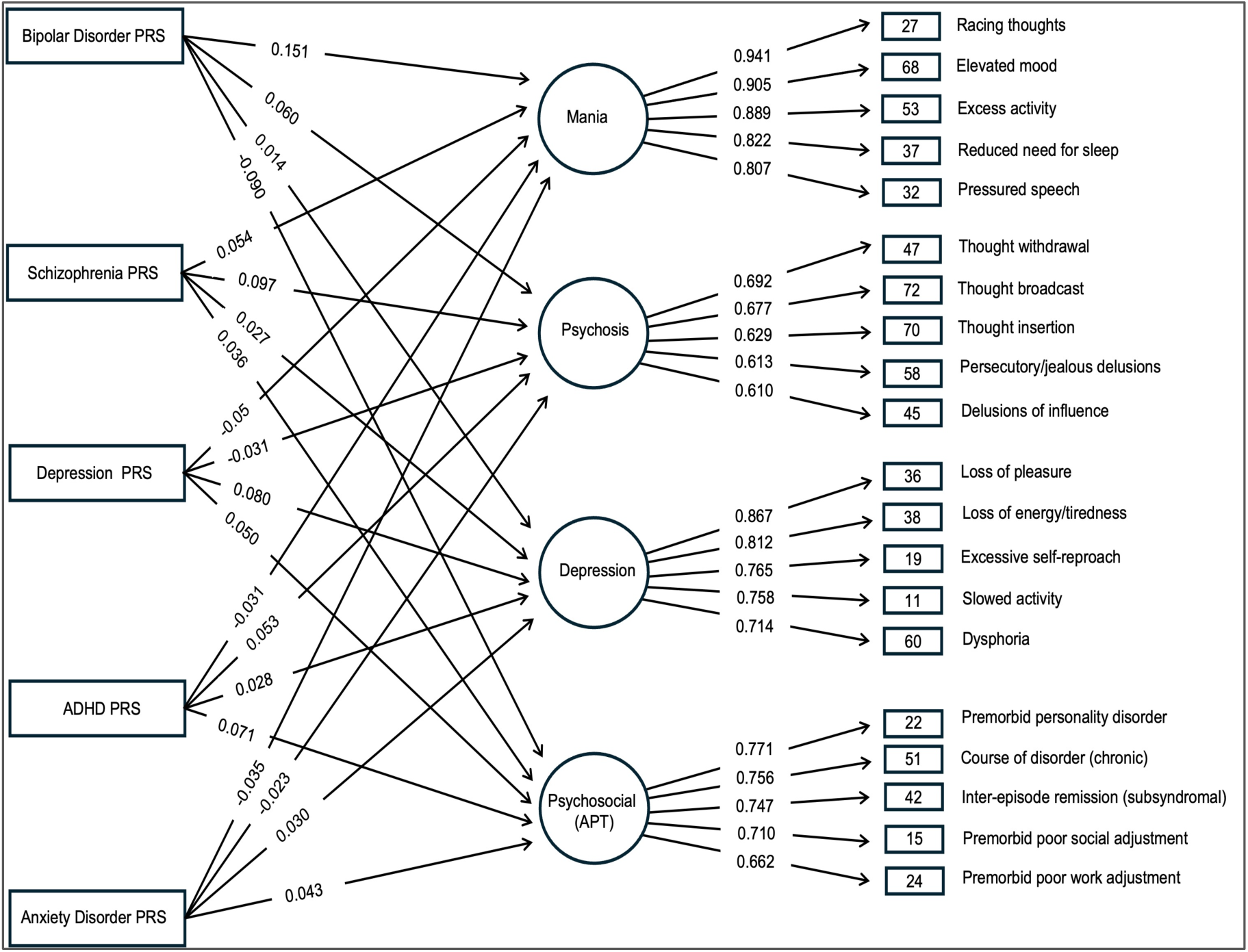
Structural equation (MIMIC) model (SEM) path diagram and fit indices. This figure illustrates the results of the Structural Equation Model (SEM) using the Multiple Indicator Multiple Cause (MIMIC) approach. Rectangles represent the five Polygenic Risk Scores (PRSs) included in the model: BD (Bipolar Disorder), SCZ (Schizophrenia), MDD (Major Depressive Disorder), ADHD (Attention-Deficit/Hyperactivity Disorder), and ANX (Anxiety). Circles indicate the four latent symptom dimensions identified through Confirmatory Factor Analysis: Mania, Psychosis, Depression, and Adverse Psychosocial Trajectory (APT). Squares represent the OPCRIT item numbers that load onto each of the four dimensions; the corresponding OPCRIT item descriptions can be found in Supplementary S9, and their Exploratory Factor Analysis (EFA) loadings are in Supplementary S7. Arrows on the left-hand side of the figure depict the associations between each of the five PRSs and the four latent symptom dimensions. Arrows on the right-hand side of the figure are representative of the associations between the latent dimensions and their respective OPCRIT items. The numerical values shown with arrows pointing between the dimension circles (on the right-hand side) indicate the covariances between the Mania dimension and the other three dimensions. The fit indices for the model are also displayed within the figure.

Items related to elevated mood loaded to the mania dimension (Regier et al., 2013). Psychotic features inclusion in BD modelling is supported by genetic liability research (Allardyce et al., 2018). Low mood/anhedonia items loaded to the depression dimension (Regier et al., 2013). Premorbid social/occupational/outcome items inclusion addressed and loaded exclusively to the APT dimension (Regier et al., 2013; Bennett et al., 2019; Judd et al., 2005). All statistical analyses were performed using R (version 4.4.2) (R Core Team, 2024).

### Sensitivity Analyses

To test EFA factor loading sensitivity, items with a cutoff of 0.4 (aligned with Allardyce et al., 2023) were later assessed. EFA item loadings (< 0.6) aligned with existing findings (Allardyce et al., 2007). Notably, Carey et al. (2024) also used a lower threshold of 0.1 for including variables in their confirmatory factor analysis. The expected correlation of rapid cycling and substance use disorders with APT dimension items is notable, as these factors are associated with poorer outcomes in BD (Lee et al., 2010; Rohde et al., 2007), potentially contributing to the lower loading reliability observed in APT (S7). Two additional sensitivity analyses assessed the robustness of symptom-dimension relationships at the individual level using factor and PRS scores (S1). All analyses were in R (version 4.4.2) with Lavaan and Bonferroni correction.

## Results

### (i) Clinical Characteristics

We examined 77 clinical symptoms and estimated five psychiatric disorder PRS in 2590 individuals with BD and 2402 healthy controls. The sample consisted of 61% females and 39% males, with no sex distribution differences across BD subtypes. A difference in age of onset was found across BD subtypes between SZA and BD2 (Table 1).

**Table 1.** Participants characteristics stratified by bipolar disorder subtypes.

| CHARACTERISTIC | OVERALL,<br>N = 2590 | SZA,<br>N = 332 | BD1,<br>N = 1475 | BD2,<br>N = 387 | BD-NOS,<br>N = 204 | P-VALUE<sup>2</sup> |
| --- | --- | --- | --- | --- | --- | --- |
| Psychosis - |  |  |  |  |  | 1.70E-77 |
| No | 34% (879 / 2590)<sup>1</sup> | 14% (47 / 332) | 36% (525 / 1475) | 79% (307 / 387) |  |  |
| Yes | 51% (1315 / 2590) | 86% (285 / 332) | 64% (950 / 1475) | 21% (80 / 387) |  |  |
| Rapid cycling - |  |  |  |  |  | 6.80E-06 |
| No | 58% (1506 / 2590) | 76% (252 / 332) | 69% (1024 / 1475) | 59% (230 / 387) |  |  |
| Yes | 27% (688 / 2590) | 24% (80 / 332) | 31% (451 / 1475) | 41% (157 / 387) |  |  |
| BD Age Onset | - | 25 (9) | 26 (11) | 28 (11) |  | 2.10E-04 |
| Sex - |  |  |  |  |  | 2.60E-01 |
| Female | 51.3% (1333 / 2590) | 59% (197 / 332) | 62% (913 / 1475) | 58% (223 / 387) |  |  |
| Male | 33.2% (861 / 2590) | 41% (135 / 332) | 38% (562 / 1475) | 42% (164 / 387) |  |  |
| Age interviewed | - | 46 (12) | 49 (13) | 51 (14) |  | 3.10E-07 |
| <sup>1</sup> Median (IQR) or Frequency (%) |  |  |  |  |  |  |
| <sup>2</sup> Kruskal-Wallis rank sum test; Pearson's Chi-squared test. |  |  |  |  |  |  |
| Abbreviations: SZA, schizoaffective disorder, BD1, bipolar disorder I, BD2, bipolar disorder II, BD-NOS, bipolar disorder not otherwise specified. |  |  |  |  |  |  |
| Note: Of the 396 participants initially categorised as 'Other', 204 were diagnosed with BD-NOS, and information was missing for the remaining 192 participants. |  |  |  |  |  |  |

### (ii) EFA

We evaluated 77 clinical symptoms in a calibration sample of 1554 BD patients (60%). Seventy-six symptoms loaded (p < 0.05) across four factors; Family history of schizophrenia (OPCRIT 13) was the exception. Symptoms exceeding 0.4 were visualised (S6-7). A four-factor EFA model fit best (χ² = 304, RMSEA = 0.033 [90% Confidence Intervals [CI] 0.024–0.037], CFI = 0.989, and TLI = 0.986). Four factors were retained based on the scree plot, parallel analysis, and lower RMSEA (S3-S5).

### (iii) CFA

We identified items with a median threshold of 0.6 for EFA factor loadings to each of four dimensions, to ensure a parsimonious CFA model with literature-relevant items. Twenty core symptoms formed a four-factor model validated by CFA. The 4-factor CFA model using 20 clinical symptoms indicated a good fit (χ² = 505.88, RMSEA = 0.03 [90% CI 0.03–0.04], CFI = 0.99, TLI = 0.99; supplementary S8). Factors, defined by highest EFA loadings, showed robust associations (p < 0.05) with all 20 symptoms. CFA generated a four-factor model with interrelated mania, psychosis, depression, and APT symptom dimensions. Lower covariances between dimensions compared to dimension items indicated distinct structures with minimal overlap.

### (iv) SEM Multiple Indicator Multiple Cause (MIMIC) model

This model indicated distinct genetic liabilities across the four clinical dimensions (Figure 1). The mania dimension, strongest associated with BD PRS, associated positively with psychosis and depression, and inversely with APT symptoms which correlated with worse outcomes. The PRS correlated strongest with their symptom dimensions; SCZ with psychosis, BD with mania, MDD with depression, and ADHD and anxiety with APT. The MIMIC (Jöreskog & Goldberger, 1975) model fit acceptably (χ² = 348.45, RMSEA = 0.04 [90% CI 0.04–0.04], CFI = 0.92, TLI = 0.90) but with less reliability than the CFA.

### (v) Sensitivity analyses

#### Individual-level factor (dimension) scores

To analyse each of the 20 core items independent contribution to the CFA model, we performed regression analyses using individual-level factor scores. Factor scores were estimated using leave-one-out CFA analyses. The median RMSEA remained relatively stable within the full CFA models confidence intervals, indicating a robust model. The 20 items were each predicted by one of the individual-level factor scores for each dimension. Participants in the top 10% of scores were more likely to report the symptom compared to those in the lower 90%. Their factor score for the symptom-related dimension was a better predictor than scores from other dimensions. The odds ratio (OR) of reporting symptoms was increased for participants in the top 10% of scores (Figure 2).

**Figure 2.**
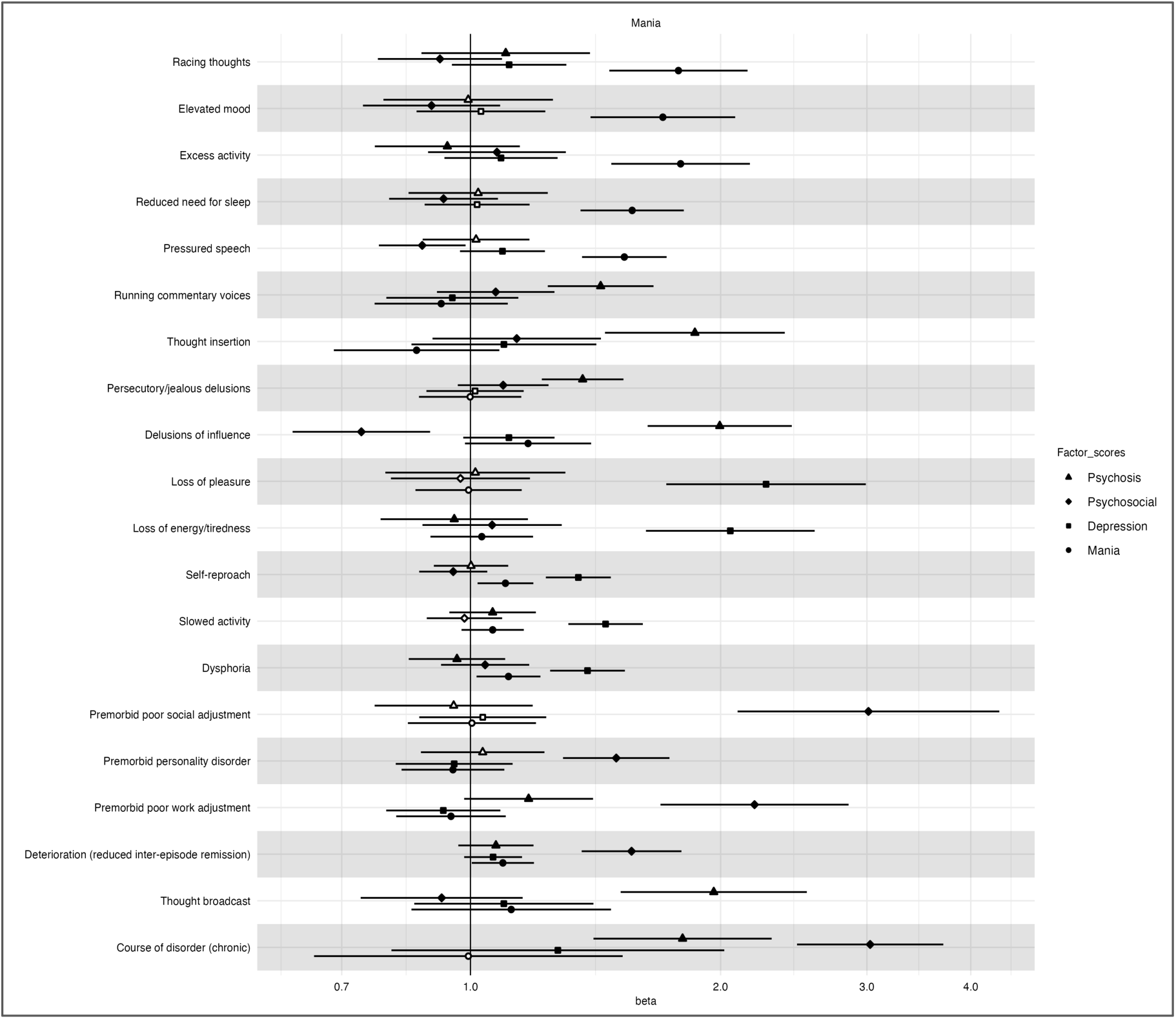
Core OPCRIT items associations using individuals leave-one-out factor scores. The odds ratio (OR) and 95% confidence intervals represent the strength of association (x-axis) between the 20 core items (y-axis) using participants factor scores for one of the four factor dimensions (individuals with the 10% scores compared to the remaining 90% of scores). Each point reflects a logistic regression model adjusted for inclusion propensity, age, age-squared and sex. An OR greater than 1 indicated an increased risk of the symptom, an OR less than 1 represented a decreased risk. The shapes denote the different (factor) dimensions. Hollow points represent tests that did not surpass the Bonferroni correction criteria.

#### Individuals PRS Scores

To ensure the five genetic contributions at the global SEM level held for each dimension item, we performed regression analyses using each item and individual-level PRS scores in turn. Participants with the top 10% compared to the lower 90% of scores for the respective dimension, were associated with a higher risk (OR) for dimension-related symptoms. Separation of global effects revealed a mixture of effect directions related to ANX and SCZ PRS for the APT dimensions (Figure 3).

**Figure 3.**
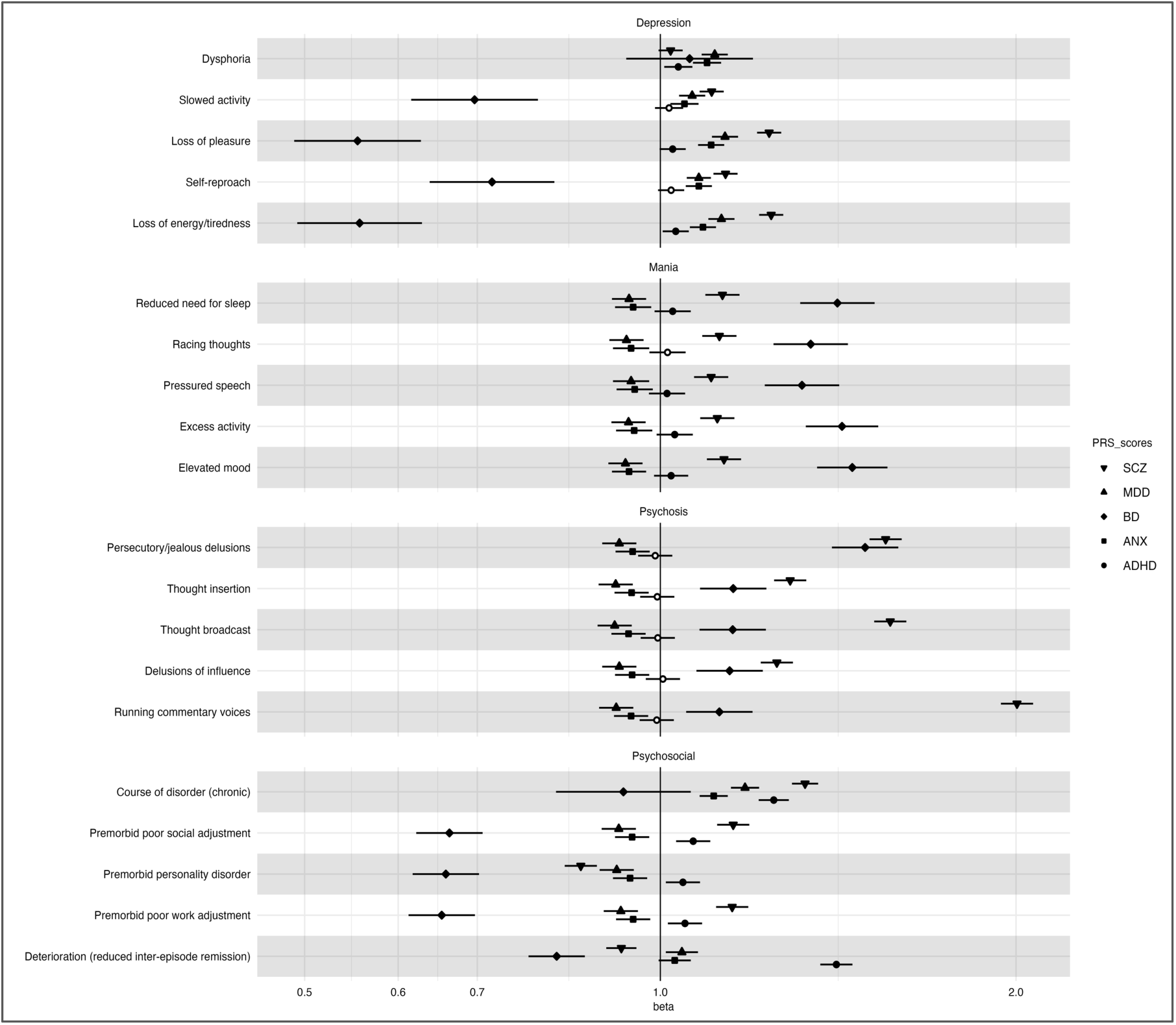
Core OPCRIT predictions using five transdiagnostic individual-level PRS scores. The odds ratio (OR) and 95% confidence intervals represent the strength of association (x-axis) between the 20 core items (y-axis) as a function of the bipolar disorder participants five PRS scores (individuals with the 10% compared to the remaining 90% of scores). Each point reflects a logistic regression model for one core item, adjusted for inclusion propensity, age, age-squared, sex, genetic ancestry and genotype array. An OR greater than 1 indicated an increased risk of the item, while an OR less than 1 represented a decreased risk. The shapes denote the different transdiagnostic PRS. Hollow points represent tests that did not surpass the Bonferroni correction criteria.

### (vi) *Post hoc* Regression Analyses

Rapid cycling (RC) considered a chronic form of BD (Coryell et al., 2003; Kupka et al., 2005), positively associated with the psychosocial dimension in EFA and inversely with mania (S7). RC also showed a positive association with premorbid social adjustment (OPCRIT 10) (OR 1.185, *P* = 1.04 × 10⁻¹⁰) and personality disorders (OR 1.391, *P* = 1.288 × 10⁻¹⁰). Premorbid personality disorder was associated with substance abuse (OPCRIT 80) (OR 1.160, *P* = 4.98 × 10⁻¹⁰) and suicidal ideation (OPCRIT 43) (OR 1.140, *P* = 1.28 × 10⁻¹⁰). Both personality disorder and RC were associated with a higher ADHD PRS (1.325 and OR 1.209, respectively, both *P* < 5 × 10⁻⁵) and an earlier onset of BD (*F* = −3.782, *P* = 6.46 × 10⁻⁶ and *F* = −3.026, *P* = 4.821 × 10⁻⁵, respectively).

## Discussion

### A Novel Four-Factor Model and the Adverse Psychosocial Trajectory Dimension

Our novel four-factor MIMIC model of BDs genetic architecture uniquely incorporates transdiagnostic genetic risk on core symptoms and an APT dimension linked to poorer outcomes. Unlike prior BD dimensional models focused on mood and psychosis (Allardyce et al., 2023), our model specifically addresses the under-examined association between premorbid psychosocial deficits and a chronic BD course.

### Genetic Links to ADHD and Anxiety

This model uniquely links a genetically influenced APT dimension, connecting premorbid deficits and adverse long-term outcomes to genetic risk for ADHD and anxiety, highlighting a distinct pathway to illness severity and their contribution to social functioning, work, personality, and a less stable BD course (Yao et al., 2023). Identifying this APT dimension and its genetic links offers a new understanding of challenges beyond BD mood episodes, suggesting a biological basis emphasising transdiagnostic risks in BDs spectrum and variable outcomes.

Our finding that a higher ADHD PRS is associated with a more adverse psychosocial trajectory (APT) in bipolar disorder aligns with evidence from Agnew-Blais et al. (2021), who demonstrated that higher ADHD genetic risk is associated with a more persistent course of ADHD into young adulthood. Supporting this, Duffy (2012) also suggests that childhood ADHD may be linked to a subtype of BD with a more severe course and poorer treatment response (Coombes et al., 2021).

Parental BD elevates child ADHD risk (Birmaher et al., 2010) and early psychosocial challenges. These factors and inherited genetic predisposition may heighten suicidality risk (McClelland, Cleare, & OConnor, 2023).

The APT dimension and its genetic links provide a new framework for understanding diverse BD clinical presentations. This highlights the need for integrated assessment and treatment, particularly when addressing co-occurring ADHD and anxiety to improve long-term psychosocial outcomes. For BD individuals with psychosocial/cognitive deficits, clinicians could tailor integrated treatment plans for optimal outcomes (McMahon, 2014).

### Dimensional Assessment and Early Intervention Implications

Our findings support dimensional assessment in BD. Evaluating an individuals psychosocial trajectory and genetic risk for associated conditions could inform more comprehensive, personalised treatment plans, suggesting earlier identification of individuals predisposed to a more challenging BD course.

This could enable preventative or early intervention strategies focused on bolstering cognitive and psychosocial functioning (Bennett et al., 2019; Krug et al., 2024). The strong genetic associations with the APT dimension, particularly with ADHD and anxiety (Duffy, 2012; Salvi et al., 2021), further underscore the potential for early intervention, as these often present in childhood and adolescence.

### Four versus a Three-Factor Model

Building on dimensional approaches, our study used more OPCRIT measures than a prior study (Allardyce et al., 2023), yielding a fourth APT dimension, validated in both our and their (eResults 4) parallel analysis. We confirmed three clinical dimensions (mania, depression, psychosis) and their genetic associations. Importantly, additional measures (S13) and PRSs loaded exclusively on our novel APT dimension, thus a four-factor model better accounts for genetic signatures in BD course specifiers. Additionally, our propensity scores adjusted for potentially inflated effect sizes.

### Predictive Utility of the PRS

Sensitivity analyses confirmed symptom strength independent of global dimensions. Factor and PRS scores better predicted risk for four dimension symptoms within than across dimensions in unseen data. PRS provided incremental predictive value to clinical data, with a median positive predictive value (PPV) at a 0.8 clinical utility threshold (S1) (Colquhoun, 2014).

### Factor Loading Thresholds

We set a 0.6 factor loading threshold for OPCRIT items due to clinical relevance and parsimony, though prior analyses used lower thresholds (Allardyce et al., 2007; Allardyce et al., 2023; Krug et al., 2024). Here, EFA robustness at 0.4 suggests future studies could use a lower threshold. While model fit and sensitivity were adequate, more items do not guarantee better accuracy and risk overfitting, reducing generalisability (Carey et al., 2024).

### Genetics of the APT Dimension

The novel APT dimension showed distinct genetic signatures. Higher BD burden indicated resilience against premorbid psychosocial deficits and chronic illness progression, predicting higher functioning in an independent BD dataset (Song et al., 2024) and an inverse relationship with rapid cycling (Coombes et al., 2020). Similarly, the mania dimension positively associated with BD PRS was inversely related to the APT dimension.

### Genetics of the APT Dimension Symptoms

Symptoms associated with BD and SCZ PRS linked to higher inter-episode remission, unlike MDD, ANX, and especially ADHD, which positively associated with reduced inter-episode remission. Higher BD PRS predicted inter-episode remission and reduced anxiety in an independent BD dataset (Song et al., 2024). Depression, anxiety, and cognitive issues are often early BD symptoms (Mignogna & Goes, 2024; Oliva et al., 2024). Rapid cycling (RC) correlated with higher ANX or ADHD PRS but inversely with BD in a prior study (Coombes et al., 2020). Co-occurring ADHD and anxiety elevate the risk for BD onset (Meier et al., 2018; Nunez et al., 2022), suggesting a less favourable trajectory.

Severe incapacity (OPCRIT 87) linked primarily to mania and psychotic features, less to APT symptoms, and least to depressive symptoms (S7). BD PRS correlated with increased symptom severity and lower depression polygenic burden in multiplex BD families (Guzman-Parra et al., 2021). Here, strongest associations existed between premorbid occupational (OPCRIT 9) and social adjustment (OCPRIT 10), and ADHD or SCZ PRS. Our finding of a negative association between APT (including premorbid adjustment) and mania aligns with the inverse relationship found by Allardyce et al. (2007). Novel to our study, is the inverse association between genetic liability to BD PRS and APT, and the inclusion of illness chronicity and personality within the APT dimension.

Longitudinal data suggests enduring psychosocial and cognitive deficits in BD (Mignogna & Goes, 2024). APT impairments affect (30-60)% of adults with BD (Bennett et al., 2019), especially with comorbid anxiety and ADHD (Salvi et al., 2021). Sensitivity analysis showed ADHD PRS consistently positive with the APT dimension symptoms, while ANX and SCZ PRS effects were more complex across indicators, suggesting nuanced relationships needing further granular investigation.

The higher BD1 proportion of cases here, linked to lower anxiety, might have limited ANX PRS and APT dimension item-level associations. Prior factor analyses found the largest BD subgroup to be characterised by affective stability with low anxiety and low risk for ADHD-like behaviours, supporting our genetic findings (Qiu et al., 2017).

ADHD PRS uniquely correlated here with a higher risk for premorbid personality disorders (OPCRIT 11) and other APT dimension symptoms. ADHD and BD comorbidity increases the risk for personality disorder and more frequent episodes, leading to poorer functioning (Miller et al., 2008). Childhood ADHD is associated with higher borderline personality disorder (BPD) risk (Miller et al., 2008).

### Limitations

Our sample, while large, primarily comprised individuals recruited through clinical settings, potentially overrepresenting those with more severe or chronic forms of BD who are more likely to seek and remain in treatment. While Inverse Probability Weighting (IPW) was applied to mitigate ascertainment, bias related to hospitalisation and symptom severity, the generalisability of our findings to community-based populations or individuals with milder presentations of BD warrants further investigation. Our decision to exclude OPCRIT items with low frequency (less than 8% missingness) could potentially limit the generalisability of our findings to individuals presenting with rarer symptoms. The cross-sectional nature of our data limits our ability to infer the temporal relationships between genetic risk, premorbid psychosocial factors, and the longitudinal course of BD. It is important to note that current PRSs for complex psychiatric disorders, including BD, explain a modest proportion of the overall variance in these conditions, and our findings, while informative at a group level, reflect trends rather than definitive individual-level predictions (Wang et al., 2022). Further research efforts, including larger genome-wide association studies and the inclusion of more diverse ancestral populations, are needed to enhance the predictive power of PRSs for clinical applications.

### Future Studies

Future research should focus on validating the four-factor models reproducibility across independent ancestral datasets, ideally utilising the same OPCRIT items to ensure comparability. Furthermore, the collection and analysis of longitudinal data will be essential for further understanding the temporal dynamics between genetic risk, the emergence of premorbid psychosocial factors, and the subsequent longitudinal course of bipolar disorder. By tracking individuals over extended periods, future studies can help to establish the precise temporal order of these events and to identify potential causal pathways. Longitudinal data incorporating detailed symptom scales could also be invaluable in identifying specific temporal links and triggers for mood episodes, especially targeting those individuals at elevated risk of suicidality.

While an individuals underlying genetic code remains relatively stable throughout their lifespan, environmental factors can influence how these genes are expressed (through epigenetic mechanisms) and interact with one another to either increase or decrease the likelihood of developing bipolar disorder. Investigating the specific mechanisms through which ADHD and anxiety might trigger or exacerbate mood episodes in BD, especially in rapid cycling, could be a logical next step, potentially involving neuroimaging or neurochemical studies to explore underlying brain circuitry.

### Conclusion

Our analysis indicates a broader transdiagnostic genetic signature, beyond traditional mood disorders, contributes to a more adverse BD trajectory, potentially worsening long-term outcomes due to psychosocial and cognitive deficits, notably linked to higher ADHD and anxiety polygenic burden.

The MIMIC model revealed a complex interplay between mania and the novel APT dimension. While ADHD PRS showed a consistent positive association with APT, ANX and SCZ PRS effects on APT items were more nuanced, requiring further research. These findings underscore the importance of considering transdiagnostic genetic risks in understanding BD heterogeneity.

### Clinical Practice

This study suggests early identification of psychosocial difficulties in individuals with higher genetic burden for ADHD and anxiety offers a crucial opportunity for interventions to improve long-term BD outcomes. Our findings underscore the potential utility of incorporating comprehensive assessments for premorbid psychosocial functioning and any co-occurring symptoms of ADHD and anxiety in individuals with or at risk for BD. This more holistic approach could facilitate the earlier identification of those individuals who may be on a more adverse psychosocial trajectory, allowing for the implementation of proactive and personalised interventions that may ultimately improve the overall course of their illness and their quality of life.

## Additional information

### Supplementary materials

Supplementary.docx

### Data availability

Data are available from AM upon reasonable request.

### Research material availability

GWAS summary statistics are publicly available.

### Analytic code availability

Code is available upon request.

### Author contribution

TV: led design and writing. AM: supported analyses. NB: revisions.

### Ethical standards

The authors assert that all procedures contributing to this work comply with the ethical standards of the relevant national and institutional committees on human experimentation and with the Helsinki Declaration of 1975, as revised in 2008. Approved by NHS MREC (MREC/03/11/090).

### Declaration of generative AI

No AI used.

## Acknowledgements

Participants data collected with support from Neuroscience Research Charitable Trust, Central London NHS Blood Transfusion Service, Camden and Islington NHS Foundation Trust, Priory Hospitals, NIHR MHRN.

## Transparency declaration

TV affirms honest, accurate, transparent account; no omissions or unexplained discrepancies.

## Author details

Correspondence email and address, Molecular Psychiatry Laboratory Division of Psychiatry University College London London, WC1E 6BT UK

