## Supplement for "Adverse Psychosocial Trajectory in Bipolar Disorder: Novel Genetic Links to ADHD and Anxiety"

### S1. Additional Methods Information

#### Controlling for Observational Study Biases

To mitigate confounders and ascertainment bias (including collider bias), which can inflate risk estimates and lead to inaccurate predictions in PRS case-only studies, this study adopted a case-control design. Including controls allows for the capture of broader genetic variation, enhancing the generalisability of findings to the wider population. Furthermore, to address potential selection bias, especially given the possibility of case selection biased toward severity, inverse-probability-weighting (IPW) (Robins, 1986) using propensity scores (van der Wal & Geskus, 2011) was applied. This adjustment aims to make the PRS more representative of the true population risk. Index bias correction was performed in R statistical software (version 4.4.2) (R Core Team, 2024) using the IPW package (van der Wal & Geskus, 2011). Stabilised weights were implemented to prevent large weights from disproportionately influencing the analysis.

#### Polygenic risk scores

Polygenic risk score profiling was performed using genotyping data from BD cases and healthy controls which was performed at the Broad Institute (Boston, MA, US) using the Affymetrix Gene 500k Assay, Illumina PsychArray, and Illumina Global Screening Array (GSA). Preimputation quality control (QC) removed variants with call rates < 0.95 (pre-filter) and < 0.98 (post-sample pruning), missing difference > 0.02, invariant positions, Minor allele frequency (MAF) < 0.01, Hardy-Weinberg equilibrium (HWE) $p < 1\times10^{-6}$ in controls and $< 1\times10^{-10}$ in cases, and samples with call rates < 0.98, FHET outside +/- 0.20, sex discrepancies, or cryptic relatedness. Variants passing QC were imputed using the Haplotype Reference Consortium (HRC) (McCarthy et al., 2016) reference panel via Eagle v2.3.5 (Loh et al., 2016) for pre-phasing and Minimac3 v2.0.1 (Das et al., 2016). The median number of post-QC variants used for association analysis was 3,164,648 (S14), utilising imputed dosage files, the European 1000 Genomes project LD reference panel (Auton et al., 2015) and principal components from PCA.

PRSs were calculated for 4992 individuals using continuous-shrinkage PRS (PRS-CS) (Ge, Chen, Ni, Feng, & Smoller, 2019) software, a Bayesian regression framework adjusting Single Nucleotide Polymorphisms (SNP) effect sizes through continuous shrinkage. The PRS-CS-auto setting was selected to learn the global shrinkage parameter from the discovery GWAS data, a pseudo-validation method comparable to grid search (Pain et al., 2021). Default settings were used for other PRS-CS parameters (Ge et al., 2019). GWAS summary statistics from the largest and most recent studies were used for BD (O'Connell et al., 2025; Bipolar Disorder Working Group of the Psychiatric Genomics Consortium) with a sample size of 840,309, depression (Howard et al., 2019; Major Depressive Disorder Working Group of the Psychiatric Genomics Consortium**)** with a sample size of 500,199, schizophrenia (Trubetskoy et al., 2022; Schizophrenia Working Group of the Psychiatric Genomics Consortium**)** with a sample size of 130,644, anxiety (Purves et al., 2020) with a sample size of 114,091 and ADHD (Demontis et al., 2023; ADHD Working Group of the Psychiatric Genomics Consortium**)** with a sample size of 225,534. These GWASs were selected as they were the largest and most recent available at the time of analysis. SCZ, MDD, ADHD and ANX are frequently comorbid with BD and share genetic risk factors, making their PRSs relevant for investigating the transdiagnostic genetic architecture of BD. PLINK v2.0 (Chang et al., 2015) score function generated raw PRS scores from posterior SNP effect means. PRSs were standardised (*M*=0, *SD*=1) in R, and power analysis was conducted using the AVENGME (Palla & Dudbridge, 2015) package in R and G*Power 3 (Faul, Erdfelder, Lang, & Buchner, 2007). Additional covariates of age at interview, sex, and genotyping array were included with IPW propensity scores, with the first 10 ancestry-specific principal components as covariates regressed out of the residual-IPW-weighted PRS scores to control for population stratification (S14-15). *P*-values were Bonferroni-corrected for multiple testing.

### Statistical analyses

#### OPCRIT items

Data for 77 clinical symptoms (S7) with adequate sample sizes were included for analysis. Balanced splits of the clinical sample were generated for use in the exploratory and subsequent confirmatory phases. Items with zero-(invariant) or near zero-variance (low frequencies) were removed to enable convergency in latent factor modelling. Items with low frequencies were subsequently interrogated separately in a series of regression analyses. Redundancy in covariates i.e., (multi)collinearity, was assessed using the function 'hetcor' in the polycor (Fox, 2022) R package to ensure parsimonious models. For items with high pairwise correlation (0.7 or above could imply multicollinearity), the item with least missingness and most clinical relevance was retained. There was low missingness (8%), 92 percent of the data was present in the 77 remaining clinical items; assessed using the 'var_miss' function in the Naniar (Tierney, Cook, & Lumley, 2023) package in R. Missingness was determined as Missingness At Random (MAR), missingness associated with other items rather than missing not at random (MNAR). This was indicated by large *p*-values when comparing items using a chi-square test via 'missing_compare' function in the finalfit package in R (finalfit team, 2023). Imputation of the data was avoided due to potential overfitting of the data (Enders, 2022). The function 'createDataPartition' in the Caret (Kuhn, 2008) package in R was used to create balanced splits of the data. The random sampling occurred within each class of subtype, stratified by sex, to preserve the overall class distribution of the data. This created two subsamples of individuals with BD, representing a 60/40 percent split in the overall clinical dataset. These 77 items were selected for analysis as they had adequate sample sizes.

#### Exploratory factor analysis

Initial exploratory factor analysis (EFA) was conducted to identify common factors from interrelated OPCRIT items using the 'efa' function in Lavaan (Rosseel, 2012) package in R. Higher factor loadings of items onto common factors, represented stronger associations between the observed variables and the underlying (latent) factor (S6-7). EFA included responses for 77 clinical symptoms (items) in 1554 (60%) of participants with a BD diagnosis, the calibration subsample (S7). The number of latent factors carried forward to confirmatory factor analysis was based on several criteria, including a Parallel Analysis (S4) using ‘fa.parallel’ function in the psych (Revelle, 2022) package in R, Scree Plot (S5) using ‘fviz_eig’ function in factoextra (Lê, Josse, & Husson, 2008) in R, fit indexes (see description of fit indices below) for models with one- to four-factors (S3) and clinical relevance. The Kaiser-Meyer-Olkin Criterion (Kaiser, 1970) (KMO) measure of sampling adequacy, i.e., how well suited the data is for EFA on a scale from 0-1, was 0.869 which was closer to 1 and well above the recommended threshold of 0.6. This indicates a proportion of the variance among variables is common variance. Bartlett’s test of sphericity (Bartlett, 1951) tested the hypothesis of unrelated variables unsuitable for structure detection. The test was significant, $p < .001$ indicating significant interrelatedness between variables which also implied the data was adequate for EFA. Clinical (scale-level) data required for Principal Component Analysis (PCA) was normalized before computing principal components, using the ‘prcomp’ function in R. Four factors were retained based on the (i) elbow (break) position in the scree plot (S5), reaffirmed in the more robust (ii) parallel analysis, and the lower RMSEA corresponding to a 4-factor model in the EFA fit indices. The threshold of 0.4 for visualising factor loadings was chosen as it aligns with criteria used in previous factor analyses of bipolar disorder symptoms, including the work by Allardyce et al. (2023). Items were ordinal categorical variables with a natural order, therefore the appropriate ‘WLSMV’ estimator was used to estimate model parameters. No differences were observed between post hoc comparison with the Maximum Likelihood (‘ML’) function to handle missing data and the pairwise deletion used by WLSMV’. Geomin factor rotation was applied to allow latent factor variables to correlate, in contrast to factor loadings which remained uncorrelated.

**Confirmatory factor analysis**

Confirmatory factor analysis was conducted to test the reproducibility of the factor loadings for 20 items within a four-factor framework, with the independent validation sample, the remaining 1036 (40%) of participants with a BD diagnosis. The selection of the 20 symptoms was based on a median factor loading above 0.6 with their relevance checked against literature on BD outcomes. This threshold of 0.6 was chosen to ensure a parsimonious and reliable model, aligning with common practices in Confirmatory Factor Analysis for selecting items that demonstrate a strong relationship with their intended latent constructs. While some guidelines suggest a higher target of 0.7, a cutoff of

0.6 is widely considered acceptable for retaining meaningful indicators of the factors. Overall model fit was assessed using the same fit indices estimated for the exploratory factor analysis. Standardised root mean squared residual (SRMR) was not reported here as elsewhere given evidence it is biased in binary datasets, as simulations data suggested (Yu & Muthén, 2002). Confirmatory factor analysis was conducted using 'cfa' function (Lavaan in R). Eigenvalues were used to compute indices for the interaction between variables, however no indices suggested multicollinearity, 'sqrt(max(ev)/value) > 15'. The linearity assumption (factor modelling) was checked by extracting factor scores using the 'lavPredict' function in lavaan in R and examined in bivariate scatterplots. The path diagrams (Figure 1, S8) were potted using lavaanPlot (Lishinski, 2024).

**Structural equation modelling with five global PRS**

To assess the five genetic contributions of each of the four latent factors identified in confirmatory factor analysis (CFA), CFA was extended to incorporate PRS predictor variables in a special case of SEM, Multiple Indicator Multiple Cause (MIMIC) model (Jöreskog & Goldberger, 1975). It is a factor model with observed covariates which explain the latent constructs in the MIMIC model. This measures both the factor as well as item-level association with genetic load in an integrated, naturalistic model for simultaneous modelling, preferable to separate multiple regression models. The analyses used the 'sem' function (lavaan in R) to implement the MIMIC model. Path diagrams, a graphical representation highlighting relationships with squares for observed variables and circles for latent factors, were generated using lavaanPlot (Lishinski, 2024). *A priori* power analysis for the SEM model was conducted using semPower (Moshagen & Bader, 2024) package in R.

**Sensitivity analyses**

**Modelling 20 core symptoms with individuals' factor scores**

A 'leave-one-out' CFA was performed to provide a more robust evaluation of the CFA model than fitting it to the entire dataset. This method assessed whether the model's stability depended on only a few specific OPCRIT items, which could affect its generalisability. The CFA was run iteratively, with each of the 20 core OPCRIT items being left out one at a time. The predictive ability of each item was then tested in unseen data. Separate logistic regression models were used for this, with the individual's factor scores (individual-level factor loadings for ‘depression’, ‘mania’, ‘psychosis’, or ‘adverse psychosocial trajectory’ (ATP) symptom dimensions) serving as predictors for the left-out item.

In each ‘leave-one-out’ CFA regression analysis, participants were randomly split into training (60%) and test (40%) sets using Caret in R. The training set underwent five-fold cross-validation to generate prediction probabilities. These probabilities were then applied to the unseen test set data to obtain predictions, evaluated using confusion matrices. Factor scores for each participant, derived from the ‘leave-one-out’ CFAs, were used in four separate regression analyses. These analyses determined the likelihood of diagnosing the left-out item, representing one of the 20 core symptoms within the four factors. The *Mdn* prediction accuracy was 0.946 (CI 95% 0.927 - 0.961, $p < .001$). The *Mdn* (IQR) positive predictive value (PPV) was 0.803 (IQR [0.774, 0.815]) approaching 80% accuracy (Colquhoun, 2014). Results were represented as odds ratios (OR) with 95% confidence intervals using the forestplot function (ggforestplot package in R (Nightingale Health, 2020)). Effects sizes, comparing the top 10% to the remaining 90% of dimension factor scores, were transformed to odds ratios (exponentiated beta coefficients). *A priori* power analysis was conducted using the pwr package in R, and *p*-values were adjusted for multiple testing using Bonferroni correction.

**Modelling 20 core symptoms with individuals’ PRS scores**

To assess the robustness of the SEM model, each of the 20 core items was tested in separate logistic regression models using the glm() function in R, with each of the five PRS (BD, SCZ, MDD, ADHD, ANX). This aimed to determine if any single item unduly influenced the correlation between a dimension and a PRS. Participants were randomly allocated to a training (60%) and a testing (40%) set using Caret. The training set was used in five-fold cross-validations to generate prediction probabilities. Predictions for the unseen test set were then obtained by combining these model probabilities with the test set data. The median prediction accuracy on the test data was 0.875 (95% CI 0.816 - 0.893), with all *p*-values below 0.001. The median (IQR) positive predictive value (PPV) was 0.769 (IQR [0.758, 0.791]). Effect sizes were presented as odds ratios with 95% confidence intervals, comparing the top 10% of PRS scores to the remaining 90%.

***Post hoc* regressions**

To test the stability of the EFA factor loadings, a sensitivity analysis was conducted using a lower cutoff of 0.4, based on the threshold applied in a previous three-factor model by Allardyce et al. (2023). This aimed to compare our findings with their earlier work and also to assess whether rapid cycling and substance use disorders, symptoms known in the literature to associate with poorer outcomes in BD, would correlate with the adverse psychosocial trajectory dimension. Correlations between rapid cycling, substance use disorders, and the adverse psychosocial trajectory dimension (S6-7) were first assessed using the 'hetcor' function in the polycor package. Effect sizes from the generalised linear models (using the glm() function in R) were transformed into odds ratios by exponentiating the beta coefficients. An *a priori* power analysis was performed for these regression models using the pwr package, and *p*-values were adjusted for multiple testing with Bonferroni correction.

All statistical analyses were carried out in R version 4.4.2 (R Core Team, 2024) on data stored securely on computer clusters supported by University College London (London, UK).

### S2. Model fit indices description

We used standard Factor Analysis (FA) fit indices for models to evaluate how well the model represented the data. The following fit indices (Moshagen & Bader, 2024) were viewed collectively to get a comprehensive view of model fit:

| | |
|---|---|
| Chi-Square Test | Represents the difference between the observed and expected covariance matrices. A non-significant *p*-value indicates a good model fit, although this test can be sensitive to sample size. |
| Comparative Fit Index (CFI) | Compares the fit of the specified model to a baseline (often a null model). Values more than .90-.95 indicate a good fit. |
| Root Mean Square Error of Approximation (RMSEA) | Measures the error of approximation in the population. Values that are lower than .05 are considered a good fit, values below .08 are considered acceptable. |
| Tucker-Lewis Index (TLI) | Like CFI, the value accounts for model complexity. A TLI above .90 suggests the model has a good fit. |

The following information in S3 to S7 below is relevant to the "Exploratory factor analysis" subsection of the Methods section and the "EFA" subsection of the Results section in the main manuscript.

#### S3. Exploratory factor analysis (EFA) models fit indices

| Model | Parameters | Chi.square | RMSEA |
|---|---|---|---|
| 1-factor | 77.00 | 877.00 | .05 |
| 2-factor | 153.00 | 678.00 | .04 |
| 3-factor | 318.00 | 552.00 | .04 |
| 4-factor | 304.00 | 462.00 | .03 |

*Note.* The data was extracted from the 1-factor to 4-factor EFA models using BD clinical symptoms.

This table (S3) presents the fit indices for Exploratory Factor Analysis (EFA) models with one to four factors, tested on a calibration subsample of bipolar disorder participants (n=1554). The fit indices included are Chi-Square, Root Mean Square Error of Approximation (RMSEA) with its 90% Confidence Intervals (CI), Comparative Fit Index (CFI), and Tucker-Lewis Index (TLI). These indices were used to evaluate the model fit for each number of factors to determine the optimal factor structure for the OPCRIT data. Lower RMSEA values and higher CFI and TLI values (typically above 0.90-0.95) generally indicate a better model fit. A four-factor EFA model fit best ($\chi^2$ = 304, RMSEA = 0.033 [90% Confidence Intervals [CI] 0.024–0.037], CFI = 0.989, and TLI = 0.986).

### S4. Parallel Analyses for exploratory factor analysis (EFA)

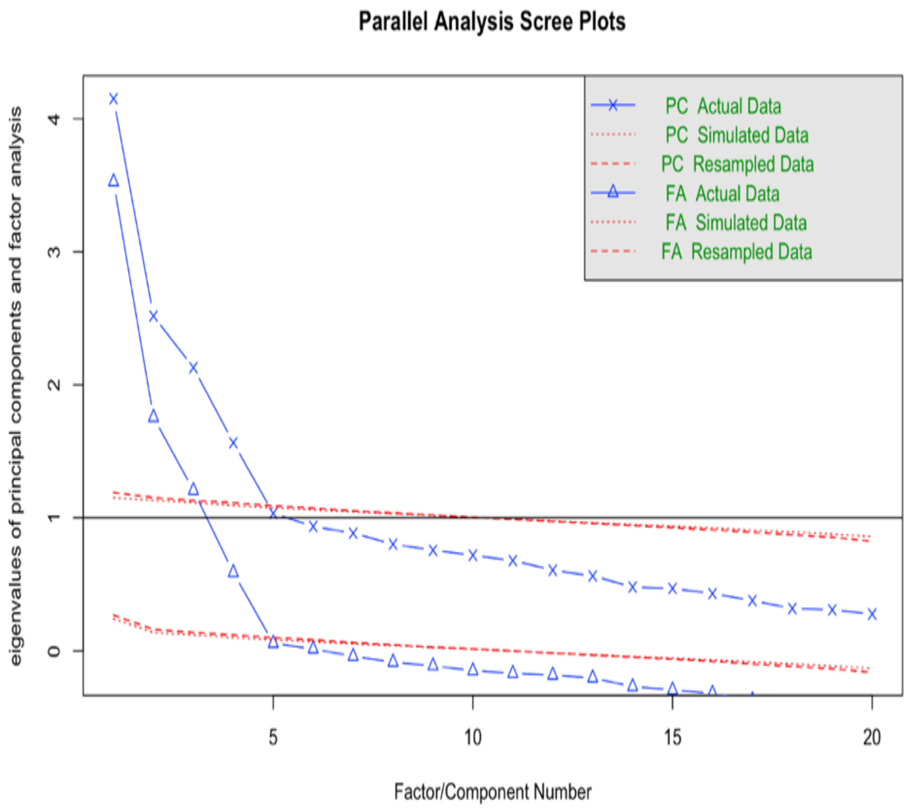

This figure (S4) illustrates the results of the parallel analysis conducted to determine the number of factors to retain in the exploratory factor analysis (EFA). The plot displays the eigenvalues obtained from the actual data (blue line) compared to the eigenvalues from random, uncorrelated data (red line). The intersection of the eigenvalues or the point where the real data eigenvalues drop below the random data eigenvalues typically suggests the appropriate number of underlying factors. In this specific analysis, the real data eigenvalues remain above the simulated data eigenvalues for four factors, suggesting that a four-factor model is appropriate. This is relevant to the "Exploratory factor analysis" subsection of the Methods section and the "EFA" subsection of the Results section in the main manuscript.

### S5. Scree plot for exploratory factor analysis (EFA)

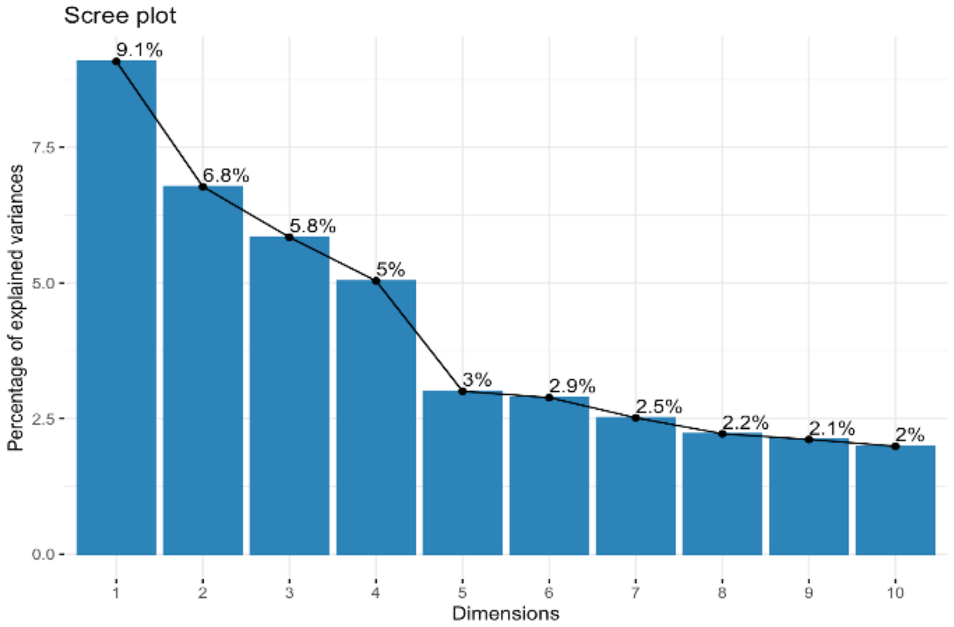

The last value of k (no. of dimensions) before the slope of the plot levels off suggests a "good" value of k.

This figure (S5) illustrates the scree plot, which is a graph of the eigenvalues of the factors plotted against the factor number. The shape of the plot helps to determine the number of factors to retain in EFA. The "elbow" or point of inflection in the scree plot typically indicates where the amount of variance explained by subsequent factors starts to diminish, suggesting an optimal number of factors before the "scree" begins. In this scree plot, the elbow is observed at the fourth factor, suggesting the retention of four factors is appropriate. This is relevant to the "Exploratory factor analysis" subsection of the Methods section and the "EFA" subsection of the Results section in the main manuscript.

### S6. Exploratory factor analysis (EFA) plot of 77 OPCRIT items

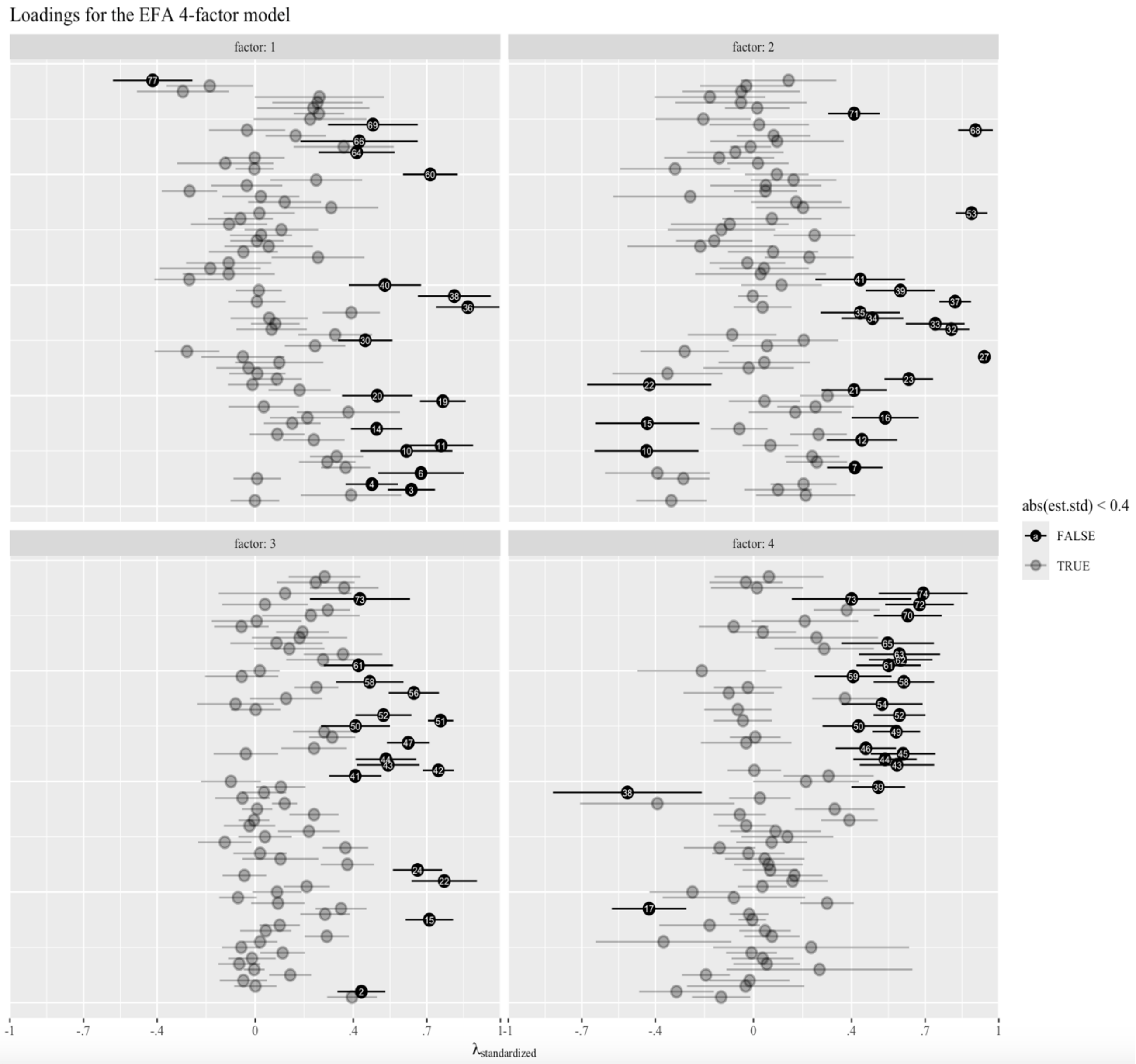

This figure (S6) visualises the standardised factor loadings (lambda values) and their 90% confidence intervals (CIs) for 77 OPCRIT items derived from Exploratory Factor Analysis. Each item is represented by a circle on the x-axis according to its factor loading. Circles are color-coded to indicate loadings above (black, ≥0.4) or below (grey, <0.4) a threshold of 0.4. The plot reveals four distinct factors, labelled as: Factor 1 - Depression, Factor 2 - Mania, Factor 3 - Adverse Psychosocial Trajectory (ATP), and Factor 4 - Psychosis. For detailed factor loading values for all items, please refer to Supplementary S7 below. This figure pertains to the "Exploratory factor analysis" subsection of the Methods section and the "EFA" subsection of the Results section in the main manuscript.

**S7. Exploratory factor analysis (EFA) loadings of 77 OPCRIT items**

Note: These 77 items were selected for analysis due to adequate sample size and less than 8% missingness.

| Item. | Item_description | Item_No. | Depres (f1) | Mania (f2) | Psychosocial (f3) | Psychosis (f4) | unique.var | communalities |
|---|---|---|---|---|---|---|---|---|
| 1 | Rapid cycling | | -0.001 | -0.335* | 0.395* | -0.132 | 0.817 | 0.183 |
| 2 | Weight loss | OPCRIT_49 | 0.391* | 0.213 | 0.433* | -0.314* | 0.435 | 0.565 |
| 3 | Diminished libido | OPCRIT_40 | 0.637* | 0.1 | 0.001 | -0.032 | 0.530 | 0.470 |
| 4 | Diurnal variation | OPCRIT_38 | 0.476* | 0.204 | -0.048 | -0.016 | 0.647 | 0.353 |
| 5 | Mode of onset | OPCRIT_5 | 0.008 | -0.286* | 0.143* | -0.194* | 0.868 | 0.132 |
| 6 | Weight gain | OPCRIT_51 | 0.676* | -0.392* | -0.004 | 0.269 | 0.263 | 0.737 |
| 7 | Early morning waking | OPCRIT_46 | 0.369* | 0.413* | -0.065 | 0.054 | 0.551 | 0.449 |
| 8 | Middle insomnia | OPCRIT_45 | 0.294* | 0.258* | -0.013 | 0.037 | 0.775 | 0.225 |
| 9 | Initial insomnia | OPCRIT_44 | 0.332* | 0.239* | 0.112 | -0.008 | 0.742 | 0.258 |
| 10 | Increased appetite | OPCRIT_50 | 0.617* | -0.436* | -0.057 | 0.235 | 0.224 | 0.776 |
| 11 | Slowed activity | OPCRIT_24 | 0.758* | 0.069 | 0.02 | -0.367* | 0.300 | 0.700 |
| 12 | Agitated activity | OPCRIT_23 | 0.239 | 0.442* | 0.292 | 0.075 | 0.532 | 0.468 |
| 13 | Stressor prior to onset | OPCRIT_16 | 0.09 | 0.265* | 0.043 | 0.047 | 0.893 | 0.107 |
| 14 | Excessive sleep | OPCRIT_47 | 0.495* | -0.058 | 0.1 | -0.179 | 0.752 | 0.248 |
| 15 | Poor premorbid social adjustment | OPCRIT_10 | 0.151 | -0.433* | 0.71* | -0.005 | 0.616 | 0.384 |
| 16 | Distractibility | OPCRIT_21 | 0.213* | 0.537* | 0.285* | -0.018 | 0.452 | 0.548 |
| 17 | Poor appetite | OPCRIT_48 | 0.38* | 0.17 | 0.35* | -0.426* | 0.477 | 0.523 |
| 18 | Inappropriate affect | OPCRIT_34 | 0.035 | 0.253* | 0.093 | 0.3* | 0.822 | 0.178 |
| 19 | Excessive self reproach | OPCRIT_42 | 0.765* | 0.045 | -0.07 | -0.08 | 0.389 | 0.611 |
| 20 | Poor concentration | OPCRIT_41 | 0.498* | 0.302* | 0.089 | -0.249 | 0.475 | 0.525 |
| 21 | Irritable mood | OPCRIT_36 | 0.181* | 0.41* | 0.21* | 0.036 | 0.668 | 0.332 |
| 22 | Premorbid personality disorder | OPCRIT_11 | -0.011 | -0.425* | 0.771* | 0.159 | 0.504 | 0.496 |
| 23 | Increased sociability | OPCRIT_53 | 0.089 | 0.633* | -0.044 | 0.167* | 0.512 | 0.488 |
| 24 | Poor premorbid work adjustment | OPCRIT_9 | 0.009 | -0.351* | 0.662* | 0.068 | 0.590 | 0.410 |
| 25 | Alcohol/drug abuse within one year of onset | OPCRIT_12 | -0.027 | -0.02 | 0.376* | 0.061 | 0.857 | 0.143 |
| 26 | Family history of schizophrenia | OPCRIT_13 | 0.098 | 0.044 | 0.103 | 0.046 | 0.969 | 0.031 |
| 27 | Thoughts racing | OPCRIT_31 | -0.05 | 0.941* | 0.02 | -0.021 | 0.150 | 0.850 |
| 28 | Unemployed | OPCRIT_7 | -0.278* | -0.281* | 0.368* | -0.138 | 0.635 | 0.365 |
| 29 | Family history of other psychiatric disorder | OPCRIT_14 | 0.244* | 0.055 | -0.124 | 0.074 | 0.904 | 0.096 |
| 30 | Restricted affect | OPCRIT_32 | 0.449* | 0.205* | 0.04 | 0.138 | 0.635 | 0.365 |
| 31 | Blunted affect | OPCRIT_33 | 0.326* | -0.087 | 0.219* | 0.09 | 0.846 | 0.154 |
| 32 | Pressured speech | OPCRIT_30 | 0.067 | 0.807* | -0.023 | -0.03 | 0.298 | 0.702 |
| 33 | Increased self esteem | OPCRIT_56 | 0.082 | 0.741* | -0.005 | 0.391* | 0.298 | 0.702 |
| 34 | Reckless activity | OPCRIT_20 | 0.057 | 0.485* | 0.240* | -0.056 | 0.657 | 0.343 |
| 35 | Relationship psychotic/affective symptoms | OPCRIT_52 | 0.392* | 0.435* | 0.008 | 0.331* | 0.364 | 0.636 |
| 36 | Loss of pleasure | OPCRIT_39 | 0.867* | 0.037 | 0.12 | -0.392 | 0.143 | 0.857 |
| 37 | Reduced need for sleep | OPCRIT_22 | 0.007 | 0.822* | -0.052 | 0.026 | 0.321 | 0.679 |
| 38 | Loss of energy/tiredness | OPCRIT_25 | 0.812* | -0.003 | 0.036 | -0.514* | 0.169 | 0.831 |

| EFA_item_No. | OPCRIT_item | OPCRIT_item_No. | Depres (f1) | Mania (f2) | Psychosocial (f3) | Psychosis (f4) | unique.var | communalities |
|---|---|---|---|---|---|---|---|---|
| 1 | Rapid cycling | | -0.001 | -0.335* | 0.395* | -0.132 | 0.817 | 0.183 |
| 2 | Weight loss | OPCRIT_49 | 0.391* | 0.213 | 0.433* | -0.314* | 0.435 | 0.565 |
| 3 | Diminished libido | OPCRIT_40 | 0.637* | 0.1 | 0.001 | -0.032 | 0.530 | 0.470 |
| 4 | Diurnal variation | OPCRIT_38 | 0.476* | 0.204 | -0.048 | -0.016 | 0.647 | 0.353 |
| 5 | Mode of onset | OPCRIT_5 | 0.008 | -0.286* | 0.143* | -0.194* | 0.868 | 0.132 |
| 6 | Weight gain | OPCRIT_51 | 0.676* | -0.392* | -0.004 | 0.269 | 0.263 | 0.737 |
| 7 | Early morning waking | OPCRIT_46 | 0.369* | 0.413* | -0.065 | 0.054 | 0.551 | 0.449 |
| 8 | Middle insomnia | OPCRIT_45 | 0.294* | 0.258* | -0.013 | 0.037 | 0.775 | 0.225 |
| 9 | Initial insomnia | OPCRIT_44 | 0.332* | 0.239* | 0.112 | -0.008 | 0.742 | 0.258 |
| 10 | Increased appetite | OPCRIT_50 | 0.617* | -0.436* | -0.057 | 0.235 | 0.224 | 0.776 |
| 11 | Slowed activity | OPCRIT_24 | 0.758* | 0.069 | 0.02 | -0.367* | 0.300 | 0.700 |
| 12 | Agitated activity | OPCRIT_23 | 0.239 | 0.442* | 0.292 | 0.075 | 0.532 | 0.468 |
| 13 | Stressor prior to onset | OPCRIT_16 | 0.09 | 0.265* | 0.043 | 0.047 | 0.893 | 0.107 |
| 14 | Excessive sleep | OPCRIT_47 | 0.495* | -0.058 | 0.1 | -0.179 | 0.752 | 0.248 |
| 15 | Poor premorbid social adjustment | OPCRIT_10 | 0.151 | -0.433* | 0.71* | -0.005 | 0.616 | 0.384 |
| 16 | Distractibility | OPCRIT_21 | 0.213* | 0.537* | 0.285* | -0.018 | 0.452 | 0.548 |
| 17 | Poor appetite | OPCRIT_48 | 0.38* | 0.17 | 0.35* | -0.426* | 0.477 | 0.523 |
| 18 | Inappropriate affect | OPCRIT_34 | 0.035 | 0.253* | 0.093 | 0.3* | 0.822 | 0.178 |
| 19 | Excessive self reproach | OPCRIT_42 | 0.765* | 0.045 | -0.07 | -0.08 | 0.389 | 0.611 |
| 20 | Poor concentration | OPCRIT_41 | 0.498* | 0.302* | 0.089 | -0.249 | 0.475 | 0.525 |
| 21 | Irritable mood | OPCRIT_36 | 0.181* | 0.41* | 0.21* | 0.036 | 0.668 | 0.332 |
| 22 | Premorbid personality disorder | OPCRIT_11 | -0.011 | -0.425* | 0.771* | 0.159 | 0.504 | 0.496 |
| 23 | Increased sociability | OPCRIT_53 | 0.089 | 0.633* | -0.044 | 0.167* | 0.512 | 0.488 |
| 24 | Poor premorbid work adjustment | OPCRIT_9 | 0.009 | -0.351* | 0.662* | 0.068 | 0.590 | 0.410 |
| 25 | Alcohol/drug abuse within one year of onset | OPCRIT_12 | -0.027 | -0.02 | 0.376* | 0.061 | 0.857 | 0.143 |
| 26 | Family history of schizophrenia | OPCRIT_13 | 0.098 | 0.044 | 0.103 | 0.046 | 0.969 | 0.031 |
| 27 | Thoughts racing | OPCRIT_31 | -0.05 | 0.941* | 0.02 | -0.021 | 0.150 | 0.850 |
| 28 | Unemployed | OPCRIT_7 | -0.278* | -0.281* | 0.368* | -0.138 | 0.635 | 0.365 |
| 29 | Family history of other psychiatric disorder | OPCRIT_14 | 0.244* | 0.055 | -0.124 | 0.074 | 0.904 | 0.096 |
| 30 | Restricted affect | OPCRIT_32 | 0.449* | 0.205* | 0.04 | 0.138 | 0.635 | 0.365 |
| 31 | Blunted affect | OPCRIT_33 | 0.326* | -0.087 | 0.219* | 0.09 | 0.846 | 0.154 |
| 32 | Pressured speech | OPCRIT_30 | 0.067 | 0.807* | -0.023 | -0.03 | 0.298 | 0.702 |
| 33 | Increased self esteem | OPCRIT_56 | 0.082 | 0.741* | -0.005 | 0.391* | 0.298 | 0.702 |
| 34 | Reckless activity | OPCRIT_20 | 0.057 | 0.485* | 0.240* | -0.056 | 0.657 | 0.343 |
| 35 | Relationship psychotic/affective symptoms | OPCRIT_52 | 0.392* | 0.435* | 0.008 | 0.331* | 0.364 | 0.636 |
| 36 | Loss of pleasure | OPCRIT_39 | 0.867* | 0.037 | 0.12 | -0.392 | 0.143 | 0.857 |
| 37 | Reduced need for sleep | OPCRIT_22 | 0.007 | 0.822* | -0.052 | 0.026 | 0.321 | 0.679 |
| 38 | Loss of energy/tiredness | OPCRIT_25 | 0.812* | -0.003 | 0.036 | -0.514* | 0.169 | 0.831 |

| | | | | | | | | |
|---|---|---|---|---|---|---|---|---|
| 39 | Grandiose Delusions | OPCRIT_57 | 0.015 | 0.599* | 0.105 | 0.509* | 0.351 | 0.649 |
| 40 | Negative formal thought disorder | OPCRIT_29 | 0.529* | 0.114 | -0.099 | 0.214 | 0.577 | 0.423 |
| 41 | Positive formal thought disorder | OPCRIT_28 | -0.269* | 0.435* | 0.408* | 0.306* | 0.587 | 0.413 |
| 42 | Inter-epsiode remission (subsyndromal) | OPCRIT_88 | -0.108 | 0.029 | 0.747* | 0.003 | 0.414 | 0.586 |
| 43 | Widespread Delusions | OPCRIT_60 | -0.183 | 0.043 | 0.542* | 0.585* | 0.373 | 0.627 |
| 44 | Well organised delusions | OPCRIT_55 | -0.108 | -0.025 | 0.533* | 0.536* | 0.440 | 0.560 |
| 45 | Delusions of influence | OPCRIT_58 | 0.256 | 0.227 | -0.038 | 0.610* | 0.435 | 0.565 |
| 46 | Other (non affective) auditory hallucinations | OPCRIT_76 | -0.048 | 0.08 | 0.24 | 0.458* | 0.732 | 0.268 |
| 47 | Life time diagnosis of cannabis abuse/depend | OPCRIT_79 | 0.055 | -0.217 | 0.624* | -0.03 | 0.398 | 0.602 |
| 48 | Single (v married) | OPCRIT_6 | 0.007 | -0.16 | 0.314* | 0.007 | 0.884 | 0.116 |
| 49 | Persecutory Delusions | OPCRIT_54 | 0.024 | 0.249* | 0.281* | 0.582* | 0.502 | 0.498 |
| 50 | Abusive/accusatory/persecutory voices | OPCRIT_75 | 0.107 | -0.131 | 0.409* | 0.428* | 0.631 | 0.369 |
| 51 | Course of disorder (chronic) | OPCRIT_90 | -0.106 | -0.097 | 0.756* | -0.043 | 0.553 | 0.447 |
| 52 | Delusions & hallucinations last for one week | OPCRIT_64 | -0.06 | 0.075 | 0.523* | 0.595* | 0.378 | 0.622 |
| 53 | Excessive activity | OPCRIT_19 | 0.017 | 0.889* | 0.002 | -0.064 | 0.191 | 0.809 |
| 54 | Primary delusional perception | OPCRIT_62 | 0.31 | 0.202 | -0.08 | 0.524* | 0.492 | 0.508 |
| 55 | Non-affective hallucination in any modality | OPCRIT_77 | 0.12 | 0.174 | 0.126 | 0.373* | 0.767 | 0.233 |
| 56 | Life time diagnosis of other abuse/depend | OPCRIT_80 | 0.023 | -0.258 | 0.647* | -0.101 | 0.397 | 0.603 |
| 58 | Persecutory/jealous delusions & hallucinations | OPCRIT_65 | -0.034 | 0.05 | 0.467* | 0.613* | 0.412 | 0.588 |
| 59 | Other primary delusions | OPCRIT_63 | 0.249 | 0.163 | -0.055 | 0.406* | 0.685 | 0.315 |
| 60 | Dysphoria | OPCRIT_37 | 0.714* | 0.095 | 0.019 | -0.211 | 0.408 | 0.592 |
| 61 | Third person auditory hallucinations | OPCRIT_73 | -0.003 | -0.320* | 0.421* | 0.551* | 0.444 | 0.556 |
| 62 | Bizarre Delusions | OPCRIT_59 | -0.122 | 0.018 | 0.277 | 0.600* | 0.572 | 0.428 |
| 63 | Running commentary voices | OPCRIT_74 | -0.002 | -0.14 | 0.358* | 0.595* | 0.511 | 0.489 |
| 64 | Delusions of guilt | OPCRIT_69 | 0.414* | -0.074 | 0.139 | 0.288* | 0.718 | 0.282 |
| 65 | Delusions of passivity | OPCRIT_61 | 0.361* | -0.012 | 0.087 | 0.547* | 0.520 | 0.480 |
| 66 | Nihilistic Delusions | OPCRIT_71 | 0.424* | 0.096 | 0.181 | 0.257 | 0.643 | 0.357 |
| 67 | Life time diagnosis of alcohol abuse/depend | OPCRIT_78 | 0.165* | 0.082 | 0.193* | 0.038 | 0.909 | 0.091 |
| 68 | Elevated mood | OPCRIT_35 | -0.033 | 0.905* | -0.056 | -0.081 | 0.205 | 0.795 |
| 69 | Delusions of poverty | OPCRIT_70 | 0.480* | 0.023 | 0.006 | 0.209 | 0.692 | 0.308 |
| 70 | Thought insertion | OPCRIT_66 | 0.224 | -0.205 | 0.227 | 0.629* | 0.476 | 0.524 |
| 71 | Impairment/incapacity during disorder | OPCRIT_87 | 0.26 | 0.41* | 0.296* | 0.38* | 0.668 | 0.332 |
| 72 | Thought broadcast | OPCRIT_68 | 0.237 | 0.015 | 0.04 | 0.677* | 0.444 | 0.556 |
| 73 | Thought echo | OPCRIT_72 | 0.254 | -0.051 | 0.427* | 0.400* | 0.577 | 0.423 |
| 74 | Thought withdrawal | OPCRIT_67 | 0.262 | -0.178 | 0.122 | 0.692* | 0.409 | 0.591 |
| 75 | Rapport difficult | OPCRIT_86 | -0.295 | -0.05 | 0.363* | 0.014 | 0.779 | 0.221 |
| 76 | Information not credible | OPCRIT_84 | -0.185 | -0.03 | 0.247* | -0.031 | 0.902 | 0.098 |
| 77 | Lack of insight | OPCRIT_85 | -0.418* | 0.143 | 0.283* | 0.063 | 0.786 | 0.214 |

The information in S8 below is relevant to the "Confirmatory factor analysis" subsection of the Methods section and the "CFA" subsection of the Results section in the main manuscript.

**S8. Confirmatory four-factor analysis path diagram (CFA) and fit indices**

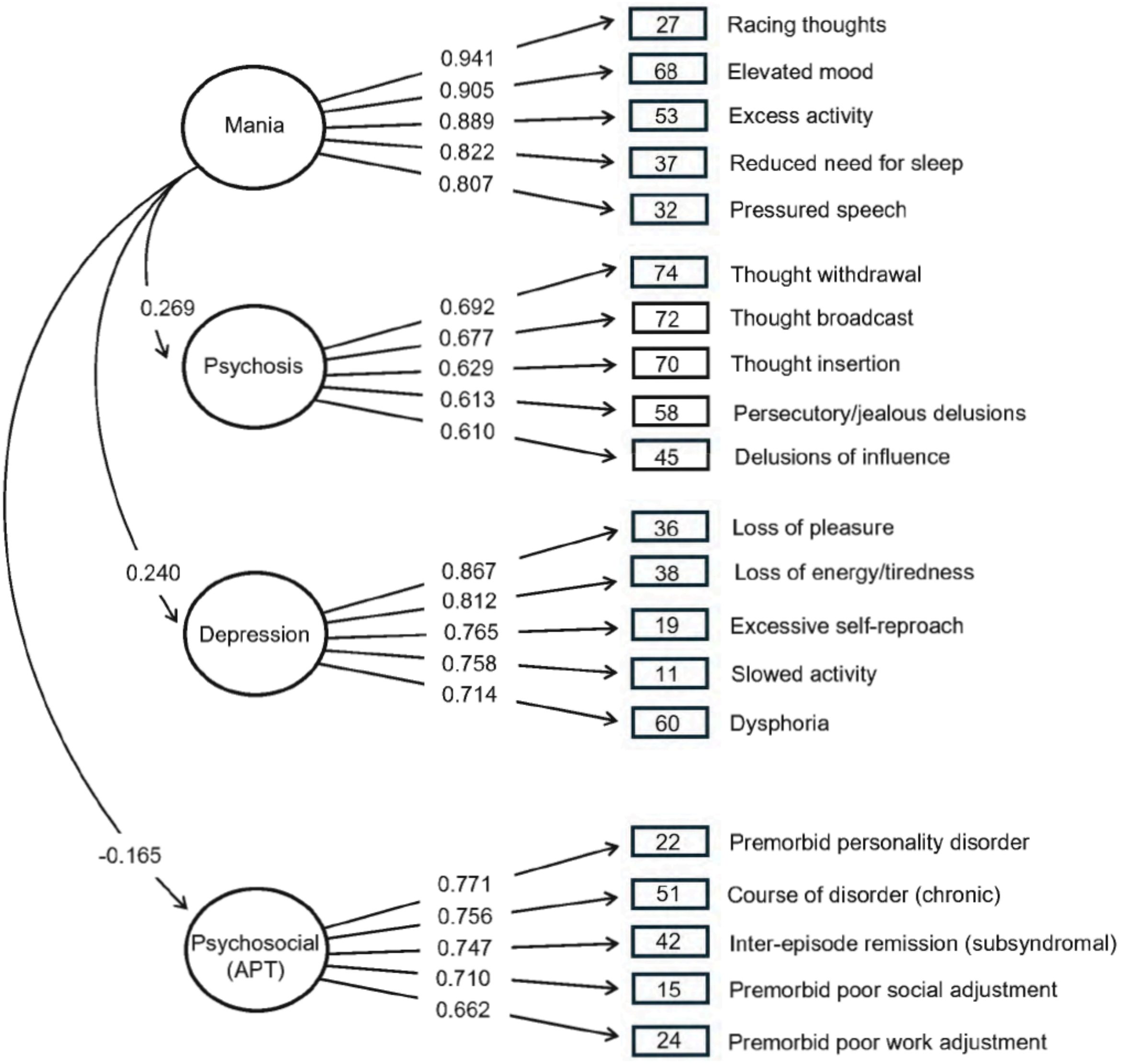

| Model | Chi.sq | RMSEA | CI.lower | CI.upper | CFI | TLI |
|---|---|---|---|---|---|---|
| 4-factor | 505.88 | .03 | 0.03 | 0.04 | .99 | .99 |

*Note.* The data was extracted from the 4-factor CFA model using 20 clinical symptoms.

This figure (S8) displays the path diagram for the four-factor Confirmatory Factor Analysis (CFA) model. The circles represent the four latent symptom dimensions: Mania, Psychosis, Depression, and

Adverse Psychosocial Trajectory (APT). The squares indicate the 20 core OPCRIT items that load onto these dimensions (refer to Supplementary S9 below for item descriptions and Supplementary S7 for Exploratory Factor Analysis loadings). Square boxes illustrate the factor loadings of each item onto its respective dimension (circle), while arrows also show the covariances between the latent dimensions. The model demonstrated good fit to the data ($\chi^2$ = 505.88, RMSEA = 0.03 [90% CI 0.03–0.04], CFI = 0.99, TLI = 0.99). Further information on the Confirmatory Factor Analysis can be found in the "Confirmatory factor analysis" subsection of the Methods section and the (iii) CFA subsection of the Results section in the main manuscript. Details regarding the model fit indices are provided in Section S2 of this supplementary material.

The following sections (S9-12) provide detailed results from the confirmatory factor analysis (CFA) and sensitivity analyses conducted to validate the four-factor model and explore the genetic contributions to the identified symptom dimensions in bipolar disorder. Specifically, these sections present the factor loadings from the CFA, and the results of the structural equation model (SEM) incorporating polygenic risk scores (PRSs), and the coefficients from analyses examining the relationship between individual OPCRIT items and both the latent factor scores and the individual PRS scores.

### S9. Confirmatory factor analysis (CFA) loadings for 20 core OPCRIT item

| Dimension | OPCRIT_item | Estimate | Std.Err | Z.value | Pvalue | ci.lower | ci.upper |
|---|---|---|---|---|---|---|---|
| Depression | Loss of pleasure (36) | 0.867 | 0.011 | 76.915 | < 0.001* | 0.809 | 0.851 |
| Depression | Loss of energy/tiredness(38) | 0.812 | 0.011 | 73.586 | < 0.001* | 0.796 | 0.839 |
| Depression | Excessive self-reproach (19) | 0.765 | 0.018 | 31.737 | < 0.001* | 0.536 | 0.607 |
| Depression | Slowed activity (11) | 0.758 | 0.015 | 43.101 | < 0.001* | 0.635 | 0.696 |
| Depression | Dysphoria (60) | 0.714 | 0.016 | 40.385 | < 0.001* | 0.615 | 0.677 |
| Mania | Racing thoughts (27) | 0.941 | 0.010 | 77.258 | < 0.001* | 0.784 | 0.825 |
| Mania | Elevated mood (68) | 0.905 | 0.011 | 73.858 | < 0.001* | 0.773 | 0.815 |
| Mania | Excess activity (53) | 0.889 | 0.010 | 80.444 | < 0.001* | 0.794 | 0.834 |
| Mania | Reduced need for sleep (37) | 0.822 | 0.012 | 60.172 | < 0.001* | 0.720 | 0.769 |
| Mania | Pressured speech (32) | 0.807 | 0.013 | 58.019 | < 0.001* | 0.711 | 0.760 |
| Psychosis | Though withdrawal (67) | 0.692 | 0.024 | 23.523 | < 0.001* | 0.516 | 0.610 |
| Psychosis | Thought broadcast (72) | 0.677 | 0.023 | 29.635 | < 0.001* | 0.650 | 0.742 |
| Psychosis | Thought insertion (70) | 0.629 | 0.025 | 20.546 | < 0.001* | 0.455 | 0.551 |
| Psychosis | Persecutory/jealous delusions (58) | 0.613 | 0.028 | 8.607 | < 0.001* | 0.184 | 0.293 |
| Psychosis | Delusions of influence (45) | 0.610 | 0.025 | 17.283 | < 0.001* | 0.389 | 0.488 |
| Psychosocial | Premorbid personality disorder (22) | 0.771 | 0.025 | 15.009 | < 0.001* | 0.331 | 0.431 |
| Psychosocial | Premorbid poor work adjustment (24) | 0.662 | 0.024 | 28.490 | < 0.001* | 0.634 | 0.728 |
| Psychosocial | Premorbid poor social adjustment (15) | 0.710 | 0.024 | 30.222 | < 0.001* | 0.684 | 0.779 |
| Psychosocial | Inter-episode remission (subsyndromal) (42) | 0.747 | 0.028 | 4.942 | < 0.001* | 0.083 | 0.192 |
| Psychosocial | Course of disorder (chronic) (51) | 0.756 | 0.026 | 12.189 | < 0.001* | 0.268 | 0.371 |
| Depression | Mania | 0.240 | 0.025 | 10.588 | < 0.001* | 0.220 | 0.320 |
| Depression | Psychosis | 0.085 | 0.031 | 2.730 | 0.006* | 0.024 | 0.146 |
| Depression | Psychosocial | 0.002 | 0.031 | 0.052 | 0.959 | -0.059 | 0.062 |
| Mania | Psychosis | 0.270 | 0.031 | 1.313 | < 0.001* | 0.209 | 0.331 |
| Mania | Psychosocial | -0.165 | 0.030 | -5.562 | < 0.001* | -0.223 | -0.107 |
| Psychosis | Psychosocial | 0.219 | 0.034 | 6.477 | < 0.001* | 0.153 | 0.285 |

This table (S9) presents the standardised factor loadings of the 20 core OPCRIT items on their respective latent dimensions (Mania, Psychosis, Depression, and Adverse Psychosocial Trajectory) derived from the confirmatory factor analysis. Significance levels for the adjusted Bonferroni *p*-values are also indicated to show the strength of the relationship between each item and its assigned dimension. This table supports the validity and internal consistency of the four-factor model. Significance levels of adjusted Bonferroni *p*-value, < 0.0001 ****, < 0.001 ***, <0.01 **, < 0.05 *

#### S10. SEM (MIMIC) of 20 OPCRIT items and five genetic covariates

| Dimension | PRS | Estimate | Std.Err | Z.value | PBonf | PBonf.signif |
|---|---|---|---|---|---|---|
| Depression | ANXprs | 0.030 | 0.028 | 1.071 | 0.041 | * |
| Depression | BDprs | 0.014 | 0.030 | 0.467 | 0.043 | * |
| Depression | ADHDprs | 0.028 | 0.029 | 0.966 | 0.333 | ns |
| Depression | MDDprs | 0.080 | 0.029 | 2.759 | 0.007 | ** |
| Depression | SCZprs | 0.027 | 0.028 | 0.964 | 0.0189 | * |
| Mania | ANXprs | -0.035 | 0.027 | -1.296 | 0.046 | * |
| Mania | BDprs | 0.151 | 0.030 | 5.033 | 5.44e-07 | **** |
| Mania | ADHDprs | -0.043 | 0.028 | -1.536 | 0.013 | * |
| Mania | MDDprs | -0.050 | 0.028 | -1.786 | 0.0086 | ** |
| Mania | SCZprs | 0.054 | 0.028 | 1.929 | 0.0035 | ** |
| Psychosis | ANXprs | -0.023 | 0.030 | -0.767 | 0.431 | ns |
| Psychosis | BDprs | 0.060 | 0.031 | 1.935 | 0.005 | ** |
| Psychosis | ADHDprs | 0.053 | 0.030 | 1.767 | 0.08 | ns |
| Psychosis | MDDprs | -0.031 | 0.030 | -1.033 | 0.306 | ns |
| Psychosis | SCZprs | 0.097 | 0.029 | 3.345 | 3e-05 | **** |
| Psychosocial | ANXprs | 0.043 | 0.033 | 1.303 | 0.003 | ** |
| Psychosocial | BDprs | -0.090 | 0.034 | -2.647 | 8e-05 | **** |
| Psychosocial | ADHDprs | 0.071 | 0.033 | 2.152 | 3e-04 | *** |
| Psychosocial | MDDprs | 0.050 | 0.033 | 1.515 | 0.003 | ** |
| Psychosocial | SCZprs | 0.036 | 0.032 | 1.125 | 0.026 | * |

This table (S10) displays the results of the Structural Equation Model (SEM) using the Multiple Indicator Multiple Cause (MIMIC) approach. It shows the path coefficients indicating the strength and direction of the relationships between the five genetic covariates (PRSs for BD, SCZ, MDD, ADHD, and ANX) and both the latent symptom dimensions and the individual 20 core OPCRIT items. Significance levels for the adjusted Bonferroni *p*-values are also included. This table illustrates the distinct genetic liabilities associated with each of the identified symptom dimensions. Significance levels of adjusted Bonferroni *p*-value, < 0.0001 ****,

< 0.001 ***, <0.01 **, < 0.05 *

**S11. Coefficients of 20 core OPCRIT items with four individual factor scores**

| OPCRIT item | Estimate | Std.Err | PBonf | Factor_scores | Dimension | PBonf.signif |
|---|---|---|---|---|---|---|
| Loss of pleasure | 0.819 | 0.141 | 2.15e-31 | Depression | Depression | **** |
| Loss of energy/tiredness | 0.720 | 0.119 | 1.45e-33 | Depression | Depression | **** |
| Self-reproach | 0.299 | 0.046 | 2.01e-38 | Depression | Depression | **** |
| Slowed activity | 0.374 | 0.053 | 2.81e-45 | Depression | Depression | **** |
| Dysphoria | 0.324 | 0.053 | 1.45e-34 | Depression | Depression | **** |
| Loss of pleasure | -0.005 | 0.075 | 9.49e-02 | Mania | Depression | ns |
| Loss of energy/tiredness | -0.031 | 0.073 | 4.94e-02 | Mania | Depression | * |
| Self-reproach | -0.097 | 0.039 | 3.34e-07 | Mania | Depression | **** |
| Slowed activity | -0.062 | 0.044 | 1.34e-03 | Mania | Depression | ** |
| Dysphoria | 0.105 | 0.045 | 1.10e-06 | Mania | Depression | **** |
| Loss of pleasure | 0.014 | 0.127 | 8.98e-02 | Psychosis | Depression | ns |
| Loss of energy/tiredness | -0.045 | 0.104 | 4.94e-02 | Psychosis | Depression | * |
| Self-reproach | 0.002 | 0.053 | 9.70e-02 | Psychosis | Depression | ns |
| Slowed activity | 0.061 | 0.061 | 8.90e-03 | Psychosis | Depression | ** |
| Dysphoria | -0.037 | 0.068 | 3.73e-02 | Psychosis | Depression | * |
| Loss of pleasure | -0.028 | 0.098 | 6.64e-02 | Psychosocial | Depression | ns |
| Loss of energy/tiredness | 0.060 | 0.098 | 3.13e-02 | Psychosocial | Depression | * |
| Self-reproach | -0.048 | 0.048 | 9.25e-03 | Psychosocial | Depression | ** |
| Slowed activity | -0.017 | 0.053 | 6.26e-02 | Psychosocial | Depression | ns |
| Dysphoria | 0.041 | 0.062 | 2.79e-02 | Psychosocial | Depression | * |

| | | | | | | |
|---|---|---|---|---|---|---|
| Racing thoughts | -0.107 | 0.081 | 1.95e-03 | Depression | Mania | ** |
| Elevated mood | -0.029 | 0.091 | 6.25e-02 | Depression | Mania | ns |
| Excess activity | -0.085 | 0.080 | 7.22e-03 | Depression | Mania | ** |
| Reduced need for sleep | -0.018 | 0.074 | 7.07e-02 | Depression | Mania | ns |
| Pressured speech | -0.089 | 0.060 | 8.66e-04 | Depression | Mania | *** |
| Racing thoughts | 0.576 | 0.098 | 2.82e-32 | Mania | Mania | **** |
| Elevated mood | 0.533 | 0.102 | 9.48e-26 | Mania | Mania | **** |
| Excess activity | 0.582 | 0.098 | 1.10e-32 | Mania | Mania | **** |
| Reduced need for sleep | 0.448 | 0.073 | 1.45e-34 | Mania | Mania | **** |
| Pressured speech | 0.426 | 0.060 | 2.81e-45 | Mania | Mania | **** |
| Racing thoughts | 0.098 | 0.119 | 1.72e-02 | Psychosis | Mania | * |
| Elevated mood | -0.006 | 0.120 | 9.64e-02 | Psychosis | Mania | ns |
| Excess activity | -0.064 | 0.103 | 3.02e-02 | Psychosis | Mania | * |
| Reduced need for sleep | 0.021 | 0.098 | 7.49e-02 | Psychosis | Mania | ns |
| Pressured speech | 0.016 | 0.075 | 7.56e-02 | Psychosis | Mania | ns |
| Racing thoughts | -0.085 | 0.088 | 1.00e-02 | Psychosocial | Mania | ** |
| Elevated mood | -0.108 | 0.097 | 5.99e-03 | Psychosocial | Mania | ** |
| Excess activity | 0.073 | 0.097 | 2.15e-02 | Psychosocial | Mania | * |
| Reduced need for sleep | -0.075 | 0.077 | 1.00e-02 | Psychosocial | Mania | ** |
| Pressured speech | -0.134 | 0.061 | 4.53e-06 | Psychosocial | Mania | **** |

| | | | | | | |
|---|---|---|---|---|---|---|
| Thought withdrawal | -0.050 | 0.093 | 3.76e-02 | Depression | Psychosis | * |
| Thought insertion | -0.093 | 0.131 | 2.45e-02 | Depression | Psychosis | * |
| Persecutory/jealous delusions | -0.013 | 0.069 | 7.79e-02 | Depression | Psychosis | ns |
| Delusions of influence | -0.107 | 0.065 | 2.85e-04 | Depression | Psychosis | *** |
| Thought broadcast | -0.093 | 0.127 | 2.30e-02 | Depression | Psychosis | * |
| Thought withdrawal | 0.081 | 0.094 | 1.51e-02 | Mania | Psychosis | * |
| Thought insertion | 0.149 | 0.117 | 2.52e-03 | Mania | Psychosis | ** |
| Persecutory/jealous delusions | 0.001 | 0.072 | 9.80e-02 | Mania | Psychosis | ns |
| Delusions of influence | 0.160 | 0.089 | 1.10e-04 | Mania | Psychosis | *** |
| Thought broadcast | 0.113 | 0.141 | 1.82e-02 | Mania | Psychosis | * |
| Thought withdrawal | 0.361 | 0.075 | 1.98e-22 | Psychosis | Psychosis | **** |
| Thought insertion | 0.622 | 0.127 | 5.76e-23 | Psychosis | Psychosis | **** |
| Persecutory/jealous delusions | 0.311 | 0.058 | 2.50e-27 | Psychosis | Psychosis | **** |
| Delusions of influence | 0.691 | 0.102 | 1.97e-41 | Psychosis | Psychosis | **** |
| Thought broadcast | 0.674 | 0.132 | 5.86e-25 | Psychosis | Psychosis | **** |
| Thought withdrawal | 0.070 | 0.083 | 1.60e-02 | Psychosocial | Psychosis | * |
| Thought insertion | 0.128 | 0.119 | 6.94e-03 | Psychosocial | Psychosis | ** |
| Persecutory/jealous delusions | 0.091 | 0.064 | 1.25e-03 | Psychosocial | Psychosis | ** |
| Delusions of influence | -0.302 | 0.097 | 2.01e-10 | Psychosocial | Psychosis | **** |
| Thought broadcast | -0.080 | 0.114 | 2.47e-02 | Psychosocial | Psychosis | * |

| | | | | | | |
|---|---|---|---|---|---|---|
| Premorbid poor social adjustment | 0.034 | 0.090 | 5.43e-02 | Depression | Psychosocial | ns |
| Premorbid personality disorder | -0.045 | 0.083 | 3.73e-02 | Depression | Psychosocial | * |
| Premorbid poor work adjustment | -0.075 | 0.081 | 1.13e-02 | Depression | Psychosocial | * |
| Inter-episode remission (subsyndromal) | 0.063 | 0.041 | 6.16e-04 | Depression | Psychosocial | *** |
| Course of disorder (chronic) | 0.242 | 0.135 | 8.10e-03 | Depression | Psychosocial | ** |
| Premorbid poor social adjustment | -0.004 | 0.091 | 9.64e-02 | Mania | Psychosocial | ns |
| Premorbid personality disorder | -0.048 | 0.073 | 2.70e-02 | Mania | Psychosocial | * |
| Premorbid poor work adjustment | -0.054 | 0.077 | 2.47e-02 | Mania | Psychosocial | * |
| Inter-episode remission (subsyndromal) | -0.090 | 0.044 | 1.46e-05 | Mania | Psychosocial | **** |
| Course of disorder (chronic) | -0.058 | 0.082 | 9.70e-03 | Mania | Psychosocial | ** |
| Premorbid poor social adjustment | -0.047 | 0.112 | 5.07e-02 | Psychosis | Psychosocial | ns |
| Premorbid personality disorder | 0.034 | 0.087 | 5.39e-02 | Psychosis | Psychosocial | ns |
| Premorbid poor work adjustment | 0.161 | 0.091 | 1.24e-04 | Psychosis | Psychosocial | *** |
| Inter-episode remission (subsyndromal) | 0.071 | 0.053 | 1.95e-03 | Psychosis | Psychosocial | ** |
| Course of disorder (chronic) | 0.588 | 0.126 | 4.60e-02 | Psychosis | Psychosocial | * |
| Premorbid poor social adjustment | 1.103 | 0.185 | 8.30e-33 | Psychosocial | Psychosocial | **** |
| Premorbid personality disorder | 0.404 | 0.075 | 3.58e-27 | Psychosocial | Psychosocial | **** |
| Premorbid poor work adjustment | 0.787 | 0.133 | 1.76e-32 | Psychosocial | Psychosocial | **** |
| Inter-episode remission (subsyndromal) | 0.446 | 0.071 | 1.59e-36 | Psychosocial | Psychosocial | **** |
| Course of disorder (chronic) | 1.108 | 0.103 | 6.96e-03 | Psychosocial | Psychosocial | ** |

This table (S11) presents the coefficients and their significance levels from the regression analyses where each of the 20 core OPCRIT items was predicted by the individual factor scores for the four latent dimensions (Mania, Psychosis, Depression, and Adverse Psychosocial Trajectory) in a 'leave-one-out' cross-validation approach. These results demonstrate the predictive ability of the factor scores for their respective symptoms in each of the four dimensions. Significance levels of adjusted Bonferroni *p*-value, < 0.0001 ****, < 0.001 ***, <0.01 **, < 0.05 *.

### S12. Coefficients of 20 core OPCRIT items with five individual PRS scores

| OPCRIT_item | Estimate | Std.Err | PBonf | PRS_score | Dimension | PBonf.signif |
|---|---|---|---|---|---|---|
| Dysphoria | 0.035 | 0.014 | 0.010 | ADHD | Depression | ** |
| Dysphoria | 0.091 | 0.014 | 0.000 | ANX | Depression | **** |
| Dysphoria | 0.057 | 0.063 | 0.000 | BD | Depression | **** |
| Dysphoria | 0.106 | 0.013 | 0.000 | MDD | Depression | **** |
| Dysphoria | 0.020 | 0.012 | 0.000 | SCZ | Depression | **** |
| Loss_of_energy/tiredness | 0.030 | 0.013 | 0.027 | ADHD | Depression | * |
| Loss_of_energy/tiredness | 0.083 | 0.013 | 0.000 | ANX | Depression | **** |
| Loss_of_energy/tiredness | -0.586 | 0.062 | 0.000 | BD | Depression | **** |
| Loss_of_energy/tiredness | 0.119 | 0.013 | 0.000 | MDD | Depression | **** |
| Loss_of_energy/tiredness | 0.216 | 0.012 | 0.000 | SCZ | Depression | **** |
| Loss_of_pleasure | 0.024 | 0.013 | 0.008 | ADHD | Depression | ** |
| Loss_of_pleasure | 0.099 | 0.013 | 0.000 | ANX | Depression | **** |
| Loss_of_pleasure | -0.590 | 0.063 | 0.000 | BD | Depression | **** |
| Loss_of_pleasure | 0.126 | 0.013 | 0.000 | MDD | Depression | **** |
| Loss_of_pleasure | 0.212 | 0.012 | 0.000 | SCZ | Depression | **** |
| Self-reproach | 0.021 | 0.013 | 0.128 | ADHD | Depression | ns |
| Self-reproach | 0.075 | 0.013 | 0.000 | ANX | Depression | **** |
| Self-reproach | -0.328 | 0.062 | 0.000 | BD | Depression | **** |
| Self-reproach | 0.075 | 0.012 | 0.000 | MDD | Depression | **** |
| Self-reproach | 0.127 | 0.012 | 0.000 | SCZ | Depression | **** |
| Slowed_activity | 0.017 | 0.014 | 0.208 | ADHD | Depression | ns |
| Slowed_activity | 0.047 | 0.014 | 0.001 | ANX | Depression | *** |
| Slowed_activity | -0.362 | 0.063 | 0.000 | BD | Depression | **** |
| Slowed_activity | 0.062 | 0.013 | 0.000 | MDD | Depression | **** |
| Slowed_activity | 0.100 | 0.012 | 0.000 | SCZ | Depression | **** |

| | | | | | | |
|---|---|---|---|---|---|---|
| Elevated_mood | 0.021 | 0.007 | 0.007 | ADHD | Mania | ** |
| Elevated_mood | -0.061 | 0.007 | 0.000 | ANX | Mania | **** |
| Elevated_mood | 0.374 | 0.035 | 0.000 | BD | Mania | **** |
| Elevated_mood | -0.068 | 0.007 | 0.000 | MDD | Mania | **** |
| Elevated_mood | 0.124 | 0.007 | 0.000 | SCZ | Mania | **** |
| Excess_activity | 0.028 | 0.008 | 0.000 | ADHD | Mania | **** |
| Excess_activity | -0.051 | 0.008 | 0.000 | ANX | Mania | **** |
| Excess_activity | 0.354 | 0.036 | 0.000 | BD | Mania | **** |
| Excess_activity | -0.062 | 0.007 | 0.000 | MDD | Mania | **** |
| Excess_activity | 0.111 | 0.007 | 0.000 | SCZ | Mania | **** |
| Pressured_speech | 0.013 | 0.008 | 0.001 | ADHD | Mania | *** |
| Pressured_speech | -0.050 | 0.008 | 0.000 | ANX | Mania | **** |
| Pressured_speech | 0.276 | 0.037 | 0.000 | BD | Mania | **** |
| Pressured_speech | -0.057 | 0.008 | 0.000 | MDD | Mania | **** |
| Pressured_speech | 0.099 | 0.007 | 0.000 | SCZ | Mania | **** |
| Racing_thoughts | 0.014 | 0.008 | 0.089 | ADHD | Mania | ns |
| Racing_thoughts | -0.057 | 0.008 | 0.000 | ANX | Mania | **** |
| Racing_thoughts | 0.293 | 0.037 | 0.000 | BD | Mania | **** |
| Racing_thoughts | -0.066 | 0.007 | 0.000 | MDD | Mania | **** |
| Racing_thoughts | 0.115 | 0.007 | 0.000 | SCZ | Mania | **** |
| Reduced_need_for_sleep | 0.024 | 0.008 | 0.003 | ADHD | Mania | ** |
| Reduced_need_for_sleep | -0.053 | 0.008 | 0.000 | ANX | Mania | **** |
| Reduced_need_for_sleep | 0.345 | 0.037 | 0.000 | BD | Mania | **** |
| Reduced_need_for_sleep | -0.061 | 0.007 | 0.000 | MDD | Mania | **** |
| Reduced_need_for_sleep | 0.121 | 0.007 | 0.000 | SCZ | Mania | **** |

| | | | | | | |
|---|---|---|---|---|---|---|
| Delusions_of_influence | 0.005 | 0.007 | 0.505 | ADHD | Psychosis | ns |
| Delusions_of_influence | -0.055 | 0.007 | 0.000 | ANX | Psychosis | **** |
| Delusions_of_influence | 0.135 | 0.033 | 0.000 | BD | Psychosis | **** |
| Delusions_of_influence | -0.080 | 0.007 | 0.000 | MDD | Psychosis | **** |
| Delusions_of_influence | 0.227 | 0.006 | 0.000 | SCZ | Psychosis | **** |
| Persecutory/jealous_delusions | -0.010 | 0.007 | 0.189 | ADHD | Psychosis | ns |
| Persecutory/jealous_delusions | -0.054 | 0.007 | 0.000 | ANX | Psychosis | **** |
| Persecutory/jealous_delusions | 0.399 | 0.033 | 0.000 | BD | Psychosis | **** |
| Persecutory/jealous_delusions | -0.080 | 0.007 | 0.000 | MDD | Psychosis | **** |
| Persecutory/jealous_delusions | 0.439 | 0.006 | 0.000 | SCZ | Psychosis | **** |
| Thought_withdrawal | -0.007 | 0.007 | 0.320 | ADHD | Psychosis | ns |
| Thought_withdrawal | -0.057 | 0.007 | 0.000 | ANX | Psychosis | **** |
| Thought_withdrawal | 0.115 | 0.033 | 0.000 | BD | Psychosis | **** |
| Thought_withdrawal | -0.086 | 0.007 | 0.000 | MDD | Psychosis | **** |
| Thought_withdrawal | 0.695 | 0.006 | 0.000 | SCZ | Psychosis | **** |
| Thought_broadcast | -0.005 | 0.007 | 0.530 | ADHD | Psychosis | ns |
| Thought_broadcast | -0.062 | 0.007 | 0.000 | ANX | Psychosis | **** |
| Thought_broadcast | 0.141 | 0.033 | 0.000 | BD | Psychosis | **** |
| Thought_broadcast | -0.089 | 0.007 | 0.000 | MDD | Psychosis | **** |
| Thought_broadcast | 0.448 | 0.006 | 0.000 | SCZ | Psychosis | **** |
| Thought_insertion | -0.006 | 0.007 | 0.429 | ADHD | Psychosis | ns |
| Thought_insertion | -0.056 | 0.007 | 0.000 | ANX | Psychosis | **** |
| Thought_insertion | 0.142 | 0.033 | 0.000 | BD | Psychosis | **** |
| Thought_insertion | -0.087 | 0.007 | 0.000 | MDD | Psychosis | **** |
| Thought_insertion | 0.253 | 0.006 | 0.000 | SCZ | Psychosis | **** |

| | | | | | | |
|---|---|---|---|---|---|---|
| Course_of_disorder_(chronic) | 0.221 | 0.015 | 0.005 | ADHD | Psychosocial | ** |
| Course_of_disorder_(chronic) | 0.104 | 0.014 | 0.000 | ANX | Psychosocial | **** |
| Course_of_disorder_(chronic) | -0.072 | 0.067 | 0.000 | BD | Psychosocial | **** |
| Course_of_disorder_(chronic) | 0.165 | 0.014 | 0.000 | MDD | Psychosocial | **** |
| Course_of_disorder_(chronic) | 0.282 | 0.013 | 0.000 | SCZ | Psychosocial | **** |
| Inter-episode_remission_(subsyndromal) | 0.343 | 0.006 | 0.007 | ADHD | Psychosocial | ** |
| Inter-episode_remission_(subsyndromal) | 0.028 | 0.006 | 0.000 | ANX | Psychosocial | **** |
| Inter-episode_remission_(subsyndromal) | -0.202 | 0.028 | 0.000 | BD | Psychosocial | **** |
| Inter-episode_remission_(subsyndromal) | 0.042 | 0.006 | 0.000 | MDD | Psychosocial | **** |
| Inter-episode_remission_(subsyndromal) | -0.076 | 0.005 | 0.000 | SCZ | Psychosocial | **** |
| Premorbid_personality_disorder | 0.044 | 0.007 | 0.006 | ADHD | Psychosocial | ** |
| Premorbid_personality_disorder | -0.059 | 0.007 | 0.000 | ANX | Psychosocial | **** |
| Premorbid_personality_disorder | -0.418 | 0.033 | 0.000 | BD | Psychosocial | **** |
| Premorbid_personality_disorder | -0.085 | 0.007 | 0.000 | MDD | Psychosocial | **** |
| Premorbid_personality_disorder | -0.155 | 0.006 | 0.000 | SCZ | Psychosocial | **** |
| Premorbid_poor_social_adjustment | 0.064 | 0.007 | 0.004 | ADHD | Psychosocial | ** |
| Premorbid_poor_social_adjustment | -0.055 | 0.007 | 0.000 | ANX | Psychosocial | **** |
| Premorbid_poor_social_adjustment | -0.411 | 0.033 | 0.000 | BD | Psychosocial | **** |
| Premorbid_poor_social_adjustment | -0.081 | 0.007 | 0.000 | MDD | Psychosocial | **** |
| Premorbid_poor_social_adjustment | 0.142 | 0.006 | 0.000 | SCZ | Psychosocial | **** |
| Premorbid_poor_work_adjustment | 0.048 | 0.007 | 0.007 | ADHD | Psychosocial | ** |
| Premorbid_poor_work_adjustment | -0.053 | 0.007 | 0.000 | ANX | Psychosocial | **** |
| Premorbid_poor_work_adjustment | -0.426 | 0.033 | 0.000 | BD | Psychosocial | **** |
| Premorbid_poor_work_adjustment | -0.077 | 0.007 | 0.000 | MDD | Psychosocial | **** |
| Premorbid_poor_work_adjustment | 0.140 | 0.006 | 0.000 | SCZ | Psychosocial | **** |

This table (S12) shows the coefficients and their significance levels from the regression analyses where each of the 20 core OPCRIT items was predicted by the five individual polygenic risk scores (BD, SCZ, MDD, ADHD, and ANX). These results illustrate the relationship between the genetic burden for each disorder and the individual clinical symptoms in each of the four dimensions. Significance levels of adjusted Bonferroni *p*-value, < 0.0001 ****, < 0.001 ***, <0.01 **, < 0.05 *

**S13. OPCRIT Items for Adverse Psychosocial Trajectory Dimension: Response Descriptions**

| Item No. | Description |
|---|---|
| 9<br>'Psychosocial (1)' | Poor work adjustment: Refers to work history before onset of illness. It should be scored if the patient was unable to keep any job for more than 6 months, had a history of frequent changes of job or was only able to sustain a job well below that expected by his educational level or training at time of first psychiatric contact. Also, score positively for a persistently very poor standard of housework (housewives) and badly failing to keep up with studies (students). (0, 1) |
| 10<br>'Psychosocial (2)' | Poor premorbid social adjustment: Patient found difficulty entering or maintaining normal social relationships, showed persistent social isolation, withdrawal or maintained solitary interests prior to onset of psychotic symptoms. (0, 1) |
| 11<br>'Psychosocial (3)' | Premorbid personality disorder: Evidence of inadequate/ schizoid/ schizotypal/ paranoid/ cyclothymic/ psychopathic/ sociopathic personality disorder present since adolescence and prior to the onset of psychotic symptoms.(0, 1) |
| 87<br>'BD outcome (4) Symptom severity' | Impairment/incapacity during disorder:<br>0 = No impairment<br>1 = Subjective impairment at work, school, or in social functioning<br>2 = Impairment in major life role with definite reduction in productivity and/or criticism has been received<br>3 = No function at all in major life role for more than 2 days or inpatient treatment has been required or active psychotic symptoms such as delusions or hallucinations have occurred |
| 88<br>'BD outcome (5) Inter-episode remission' | Deterioration from premorbid level of functioning: Patient does not regain his premorbid social, occupational or emotional functioning after an acute episode of illness. (0, 1) |
| 90<br>'BD outcome (6) Illness recovery to chronic course' | Course of disorder:<br>1 = Single episode with good recovery<br>2 = Multiple episodes with good recovery between<br>3 = Multiple episodes with partial recovery between<br>4 = Continuous chronic illness<br>5 = Continuous chronic illness with deterioration<br>(nb score this item in hierarchical fashion, e.g. if patient's course in past rated '2',but for the time-period now being considered it rates '4', then the correct rating is '4'.) (1, 2, 3, 4, 5,) |
| Note | OPCRIT (version 4) (Williams et al., 1996) includes 90 items of psychopathology, premorbid functioning, personal and family history information. Expansion of OPCRIT necessitated an increase in the number of items comprising the OPCRIT checklist beyond the original 74-item checklist. Lifetime occurrence was assessed for each patient. Inter-rater reliability was formally assessed using 20 randomly selected cases (mean κ Statistic = 0.85). |

### S14. Frequency of genotype array across participants

| Sample | Affymetrix Gene Chip 500k | Illumina Global Screening Array | Illumina PsychChip | *N* |
|---|---|---|---|---|
| Post-QC'd Sample size | 491 cases, 495 controls | 416 cases, 533 controls | 1683 cases, 1374 controls | 4992 |
| Percent % | 0.197 | 0.190 | 0.613 | |
| Post-QC'd Imputed SNPs | 3,080,075 | 3,164,648 | 3,443,778 | *Mdn*=3,164,648 |

To address potential confounding due to population stratification and study inclusion biases, Inverse Probability Weighting (IPW) was applied (age at interview, sex, genotyping array (S14)), and population stratification was controlled for by regressing out the first 10 ancestry-specific principal components from the IPW-weighted polygenic risk scores for all disorders (BD, SCZ, ADHD, MDD, and anxiety). For illustrative purposes, the pre- and post-correction ANOVA results for the BD PRS are presented in S15. The application of this correction had a similar direction and magnitude of effect on the association of the other PRSs, attenuating the uncorrected associations.

### S15. BD PRS pre- and post correction for covariates (IPW)

| BD PRS | Analysis of Variance (ANOVA) | | |
|---|---|---|---|
| | *F* | *df* | *P* |
| Pre-correction | 85.22 | 2, 4989 | p < .001 |
| Post-correction | .092 | 2, 4989 | .912 |
